# Daily Versus Intermittent Oral Iron Supplementation for the Treatment of Anaemia in Low- and Middle-Income Countries: A Systematic Review and Meta-analysis

**DOI:** 10.64898/2026.07.21.26358553

**Authors:** Pratibha Dhiman, Mrunali Zode, Ranadip Chowdhary, Pravin Kumar, Subhanwita Manna, Devoleena Chatterjee, Arshi Ahad, Krushna Chandra Sahoo, Tanica Lyngdoh, Reema Mukherjee

## Abstract

**Background:** Iron deficiency anaemia remains a major public health problem in low- and middle-income countries (LMICs), particularly among children, adolescents, women of reproductive age, and pregnant women. Although daily oral iron supplementation is the standard treatment, uncertainty remains regarding whether intermittent dosing provides comparable efficacy with better tolerability and acceptability. We conducted a systematic review and meta-analysis to compare the effectiveness and safety of daily versus intermittent oral iron or iron-folic acid supplementation for treating anaemia in LMICs.

**Methods:** Four electronic databases (PubMed/MEDLINE, Embase, Scopus, and Web of Science) were searched from inception to July 2025. Randomised controlled trials, quasi-experimental studies, and prospective cohort studies conducted in LMICs among anaemic children, adolescents, women of reproductive age, and pregnant women were included. Primary outcomes were changes in haemoglobin concentration and serum ferritin from baseline to study end. Secondary outcomes included adherence, adverse effects, anaemia recovery, and maternal and fetal outcomes. Random-effects meta-analysis, subgroup analysis, and GRADE certainty assessment were performed.

**Results:** Fourteen studies met the inclusion criteria, with 13 studies (19 comparisons; 724 participants receiving daily supplementation and 948 receiving intermittent supplementation) included in the meta-analysis. Daily iron supplementation was associated with a small but statistically significant increase in haemoglobin compared with intermittent regimens (MD=0.34 g/dL; 95% CI: 0.08, 0.60; I²=87.8%). Subgroup analysis demonstrated benefits among children (MD=0.44 g/dL; 95% CI: 0.19, 0.68; I²=68.1%) and pregnant women (MD=0.79 g/dL; 95% CI: 0.04, 1.55; I²=87.8%), whereas no significant difference was observed among adolescents. Daily supplementation also resulted in higher serum ferritin concentrations (MD=4.48 µg/L; 95% CI: 0.37, 8.59). Intermittent regimens were associated with higher adherence and fewer gastrointestinal adverse effects.

**Conclusion:** Very low-certainty evidence suggests daily supplementation may produce a small increase in haemoglobin and serum ferritin compared with intermittent regimens, although the magnitude and clinical significance of these differences remain uncertain. Intermittent regimens were associated with fewer gastrointestinal side effects and better adherence. Where adherence or treatment tolerability is a concern, intermittent regimens may represent a pragmatic alternative to daily supplementation, particularly in resource-constrained LMIC settings.

## INTRODUCTION

Anaemia remains a significant global public health burden, affecting an estimated 1.92 billion worldwide, with overall prevalence of 24.3% (1). Iron deficiency is the leading cause of anaemia, accounting for nearly 85% of anaemia-related years lived with disability globally (1). The burden is concentrated in low- and middle-income countries (LMICs), particularly in South Asia and sub-Saharan Africa, where women of reproductive age, pregnant women, adolescents and young children are disproportionately affected (1,2). In LMICs, anaemia affects approximately 40% of women of reproductive age, 45% of pregnant women and more than half of children (6 months-9 years) years of age (3–5). India bears one of the highest burdens globally, with anaemia affecting 57% of women of reproductive age, 52% of pregnant women, 59% of adolescent girls and 67% of children according to the National Family Health Survey-5 (6,7). The consequences of untreated iron deficiency anaemia (IDA), characterized by reduced haemoglobin concentrations, include impaired cognitive development, reduced physical capacity, adverse pregnancy outcomes, and increased susceptibility to infection, collectively contributing substantially to morbidity, reduced productivity, and loss of human capital in high-burden settings (8).

Oral iron supplementation is the recommended first-line treatment for iron deficiency anaemia, and most national and international guidelines recommend daily elemental iron therapy (9,10). However, adherence to daily treatment remains poor, with studies consistently reporting substantial non-compliance, largely because of gastrointestinal side effects, forgetfulness, and the prolonged duration of therapy (11,12). These challenges are particularly important in LMICs, where anaemia is highly prevalent and dietary iron intake is often inadequate (1,8).

Consequently, intermittent oral iron supplementation has been proposed as a potential alternative to improve adherence while maintaining treatment efficacy (13,14). The rationale for intermittent supplementation is supported by current understanding of iron homeostasis. Hepcidin, the principal regulator of iron metabolism, increases after oral iron administration and suppresses intestinal iron absorption for approximately 24 hours. Consequently, daily dosing may maintain persistently elevated hepcidin concentrations, limiting absorption of subsequent doses. In contrast, alternate-day or intermittent dosing allows hepcidin concentrations to decline between doses, potentially improving fractional iron absorption (15). A randomised trial demonstrated greater iron absorption with 60 mg elemental iron administered on alternate days than with consecutive daily dosing (16). In addition, administering iron less frequently may reduce the accumulation of unabsorbed iron in the gastrointestinal tract, thereby decreasing gastrointestinal adverse effects and improving adherence (11,13,14).

However, evidence comparing daily and intermittent oral iron supplementation remains inconclusive. Several systematic reviews and meta-analyses have examined this question, but important evidence gaps remain. Many reviews have primarily evaluated intermittent iron supplementation for the prevention of anaemia rather than its use in the treatment of established anaemia (13,17). Furthermore, existing reviews have generally focused on individual population groups, including children, pregnant women, or menstruating women, thereby limiting the generalisability of their findings across the broader spectrum of anaemic populations (13,17–20). Several reviews have also combined evidence from high-income countries and LMICs, despite substantial differences in dietary iron intake, baseline iron deficiency, infectious disease burden, healthcare delivery, and treatment adherence, potentially limiting the applicability of pooled findings to resource-constrained settings (8,14,17). To our knowledge, no systematic review has comprehensively synthesized evidence on the comparative effectiveness and safety of daily versus intermittent oral iron supplementation exclusively for the treatment of anaemia across multiple high-risk populations in LMICs. Given the high burden of anaemia and the persistent challenge of poor adherence to daily oral iron supplementation in LMICs, it is important to determine whether intermittent dosing provides similar therapeutic effectiveness while improving treatment adherence and tolerability.

Therefore, we conducted a systematic review and meta-analysis to compare daily versus intermittent oral iron or iron-folic acid supplementation for the treatment of anaemia among children, adolescents, women of reproductive age, and pregnant women in LMICs. The primary outcomes were changes in haemoglobin concentration and serum ferritin concentrations. Secondary outcomes included adherence, gastrointestinal adverse effects, recovery from anaemia, maternal and fetal outcomes where applicable, and other reported clinical outcomes.

## METHODS

The protocol was registered with the Open Science Framework (OSF) (https://doi.org/10.17605/OSF.IO/EFWTV). This review was conducted and reported in accordance with the Preferred Reporting Items for Systematic Reviews and Meta-Analyses (PRISMA) 2020 guideline (21). No ethical approval was required as only published aggregate data were used.

### Eligibility criteria Inclusion criteria

We included studies that (1) enrolled anaemic participants living in LMICs (According to the World Bank classification), comprising children (6 months-9 years), adolescents (10-19 years), women of reproductive age (15-49 years), and pregnant women; (2) compared daily oral iron or iron-folic acid (IFA) supplementation with an intermittent regimen (alternate-day, twice-weekly, or weekly); (3) reported haemoglobin and/or serum ferritin concentrations; and (4) were randomised controlled trials (RCTs), quasi-experimental studies, or prospective cohort studies with a concurrent comparison group.

### Exclusion criteria

We excluded (1) studies conducted in high-income countries; (2) studies enrolling non-anaemic participants; (3) trials in which iron or IFA was co-administered with additional nutritional supplements that differed between study groups; and (4) conference abstracts, editorials, letters, and review articles.

### Information sources and search strategy

Four electronic databases (PubMed/MEDLINE, Embase, Scopus, and Web of Science) were searched from database inception to July 2025. The search strategy combined Medical Subject Headings (MeSH) and free-text terms describing the population (children, adolescents, pregnant women, women of reproductive age), intervention and comparator (oral iron supplementation, IFA, ferrous sulphate, daily, intermittent, alternate-day, weekly), outcomes (haemoglobin, serum ferritin, adverse effects), and setting (LMICs and developing countries). The complete search strategy is provided in Supplementary Table 1. Reference lists of all included studies and relevant systematic reviews were searched to identify additional eligible studies.

### Study selection

All retrieved citations were imported into Rayyan for duplicate removal and study screening (22). Reviewers (PD, MZ, SM, PK) independently screened titles and abstracts, followed by full-text assessment of potentially eligible studies. Disagreements were resolved through discussion or, when necessary, by consultation with a third reviewer (RM). The study selection process is summarised in the PRISMA 2020 flow diagram.

### Outcome measures

The primary outcomes were changes in haemoglobin concentration (g/dL) and serum ferritin concentration (µg/L) from baseline to the end of the intervention. For each study, mean change from baseline (endline minus baseline) was calculated separately for the daily and intermittent supplementation groups, and the difference in mean changes between groups was pooled using the mean difference (MD) to estimate comparative treatment effectiveness. These outcomes reflected the comparative effectiveness of daily versus intermittent iron or IFA supplementation on haematological parameters.

Secondary outcomes included recovery from anaemia, maternal and infant outcomes (birth weight, preterm birth, and maternal anaemia at delivery), adherence, adverse effects (including gastrointestinal symptoms, nausea, and constipation), and long-term outcomes such as anaemia recurrence, growth, and functional status. All outcomes were pre-specified before data extraction; outcomes not reported by individual studies were recorded as unavailable.

### Data extraction

Two reviewers independently extracted data using a pre-piloted, standardized Microsoft Excel data extraction form. Extracted variables included study characteristics (author, publication year, country, study design, and setting), participant characteristics (sample size, age, sex, and baseline and endline haemoglobin concentration), intervention and comparator details (iron formulation, elemental iron dose, dosing frequency, and duration), and outcome data (means, standard deviations [SDs], standard errors [SEs], and change scores for haemoglobin and serum ferritin).

For studies reporting SEs, SDs were calculated using SD = SE × √n. Where change scores and corresponding SDs were not reported, they were estimated from baseline and endline means and SDs assuming a correlation coefficient of 0.5 between repeated measurements, consistent with recommendations in the Cochrane Handbook (23).

### Risk of bias assessment

Risk of bias in RCTs was assessed using the Cochrane Risk of Bias 2·0 (RoB 2) tool across five domains: randomisation process, deviations from intended interventions, missing outcome data, measurement of the outcome, and selection of the reported result. Non-randomised studies were assessed using ROBINS-I. Two reviewers (PD and MZ) independently assessed each study; disagreements were resolved through third-reviewer adjudication.

### Statistical analysis

Meta-analysis was performed using Stata version 18 (StataCorp, College Station, TX, USA). A random-effects model using restricted maximum likelihood (REML) estimation was employed because clinical and methodological heterogeneity across studies was anticipated, consistent with recommendations in the Cochrane Handbook for Systematic Reviews of Interventions (23). Treatment effects were summarised as mean differences (MDs) with 95% confidence intervals (CIs). Statistical heterogeneity was assessed using Cochran’s Q statistic, I², and τ².

The primary analysis included all eligible studies. Analyses were conducted according to population groups (children, adolescents, women of reproductive age, and pregnant women).

Publication bias was assessed by visual inspection of funnel plots and Egger’s regression test. A sensitivity analysis was performed by sequentially excluding individual studies to evaluate the robustness of the pooled estimates. The certainty of evidence for the primary outcomes was assessed using GRADE approach.

## Results

### Study Selection and Characteristics

Systematic searches across four databases retrieved 6,444 records. After removing 2,530 duplicates, 3,914 records underwent title and abstract screening, of which 75 full-text articles were assessed for eligibility. Ultimately, 14 studies met the inclusion criteria and were included in the review, comprising both quantitative (meta-analysis) and narrative syntheses. Studies were primarily excluded because they evaluated multi-micronutrient interventions, focused on food fortification, enrolled non-anaemic populations, or were conducted in high income settings. The study selection process is presented in Figure 1.

**Figure 1:**
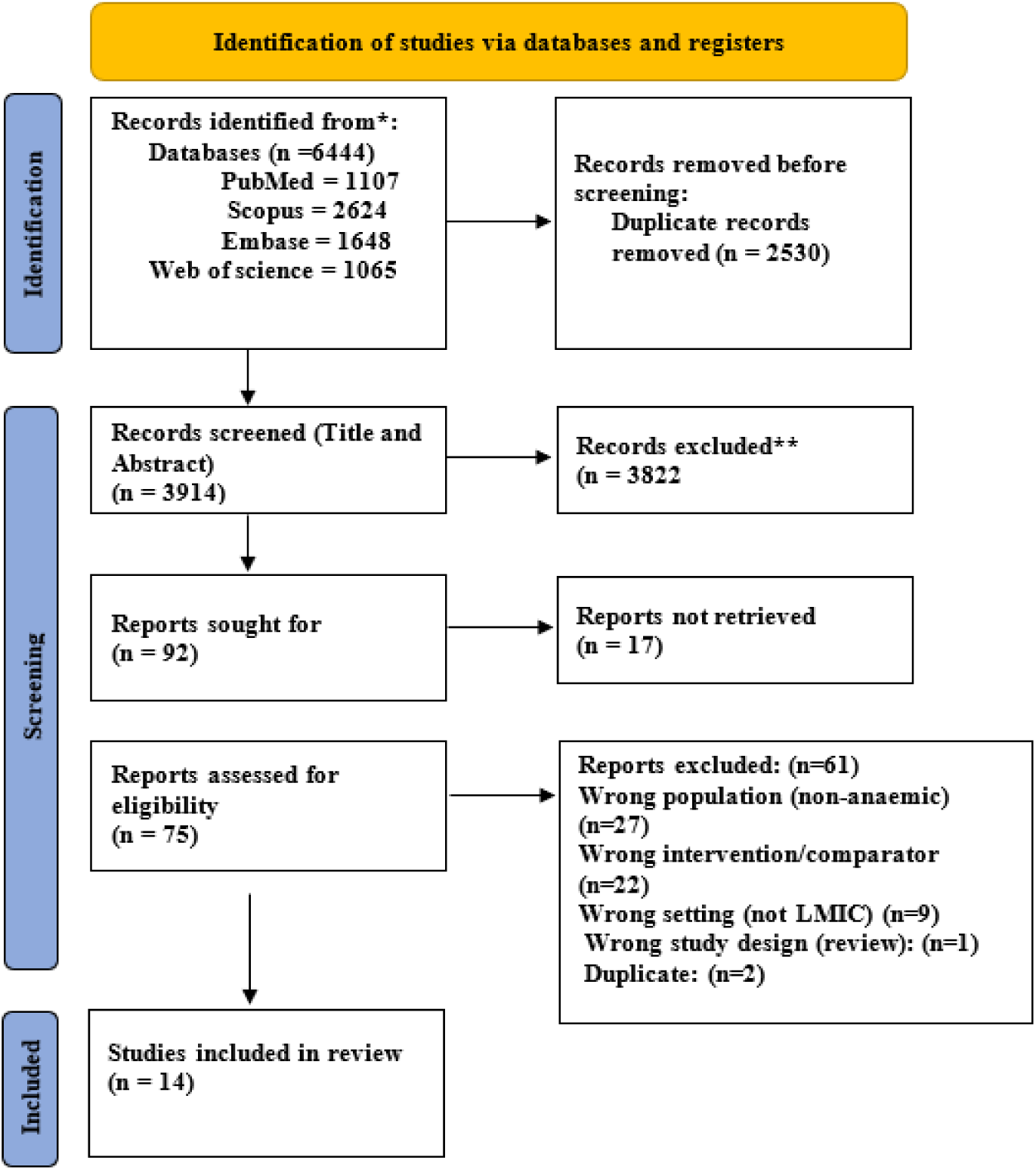
PRISMA 2020 flow diagram for new systematic reviews which included searches of databases and registers only.

A total of 14 studies comprising 20 comparisons were included in this review (24–37). Of these, 13 studies (19 comparisons) were included in the meta-analysis, comprising 724 participants allocated to daily supplementation and 948 participants allocated to intermittent supplementation (24–36). One study by Kaundal et al. (37) was excluded from the meta-analysis because outcomes were reported as medians without corresponding means and standard deviations, precluding quantitative synthesis. Its findings were therefore summarised narratively. The included studies comprised both randomised and non-randomised intervention designs. The majority were randomised controlled trials (RCTs), while two studies used non-randomised designs (Table 1).

**Table 1.**
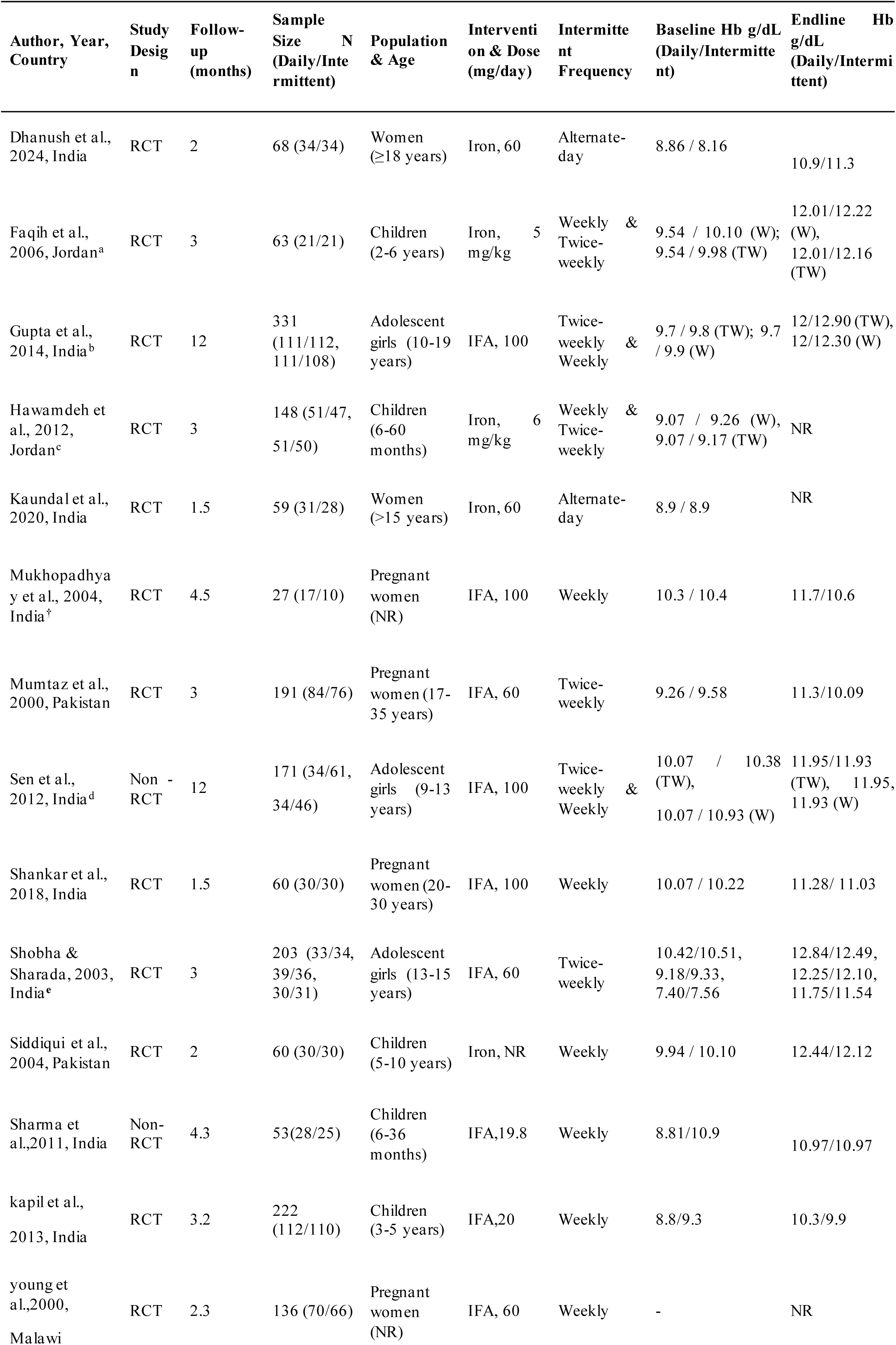
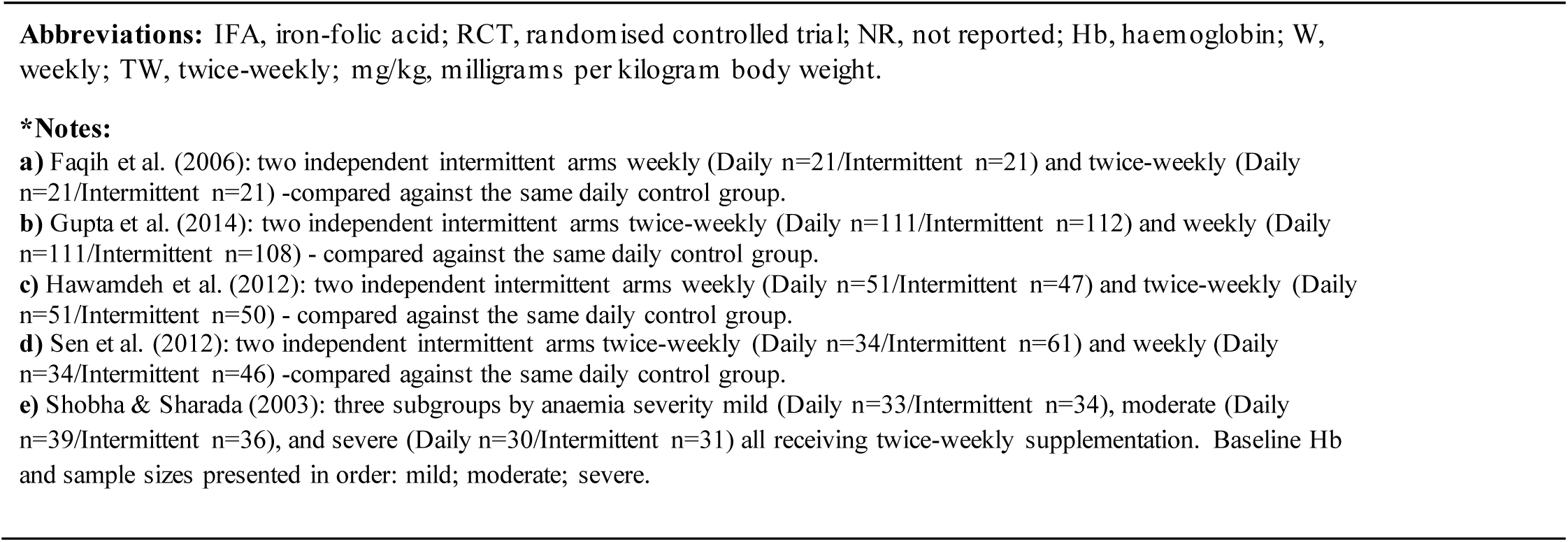
Characteristics of Included Studies.

Participants ranged from preschool children to pregnant women, with baseline mean haemoglobin concentrations generally ranging from approximately 8 to 10 g/dL across studies. Daily elemental iron doses varied from 5 to 100 mg, whereas intermittent regimens typically administered 30-120 mg once or twice weekly (Table 1). Intervention duration ranged from 4 weeks to 12 months. Study characteristics, participant populations, intervention regimens, and comparator details are summarised in Table 1. All included studies reported the primary outcome of change in haemoglobin concentration, while five studies assessed serum ferritin concentrations (25–27,31,34)). Six studies reported recovery from anaemia (25,27,30,32,34,36), two reported maternal and/or fetal outcomes (28,31), six assessed treatment adherences (24,26,28,32,34,37), and five reported adverse events, primarily gastrointestinal side effects (24,26,28,32,37). None of the included studies reported long-term clinical outcomes, such as anaemia recurrence, growth, or functional status.

### Primary outcome

#### Pooled change in haemoglobin concentration

The meta-analysis compared changes in haemoglobin concentration between daily and intermittent oral iron or iron-folic acid supplementation across the included studies. Thirteen studies contributing 19 comparisons were included in the meta-analysis, comprising 724 participants receiving daily supplementation and 948 receiving intermittent supplementation. Using a random- effects model, the pooled mean difference (MD) in haemoglobin change was 0.34 g/dL (95% CI 0.08 to 0.60), favouring daily supplementation. Although the magnitude of the effect was modest, the overall difference was statistically significant (Z = 2.60, p = 0.01).

There was substantial between-study heterogeneity (I² = 87.8%; τ² = 0.28; p < 0.001), indicating considerable variability in treatment effects across studies (Figure 2).

**Figure 2.**
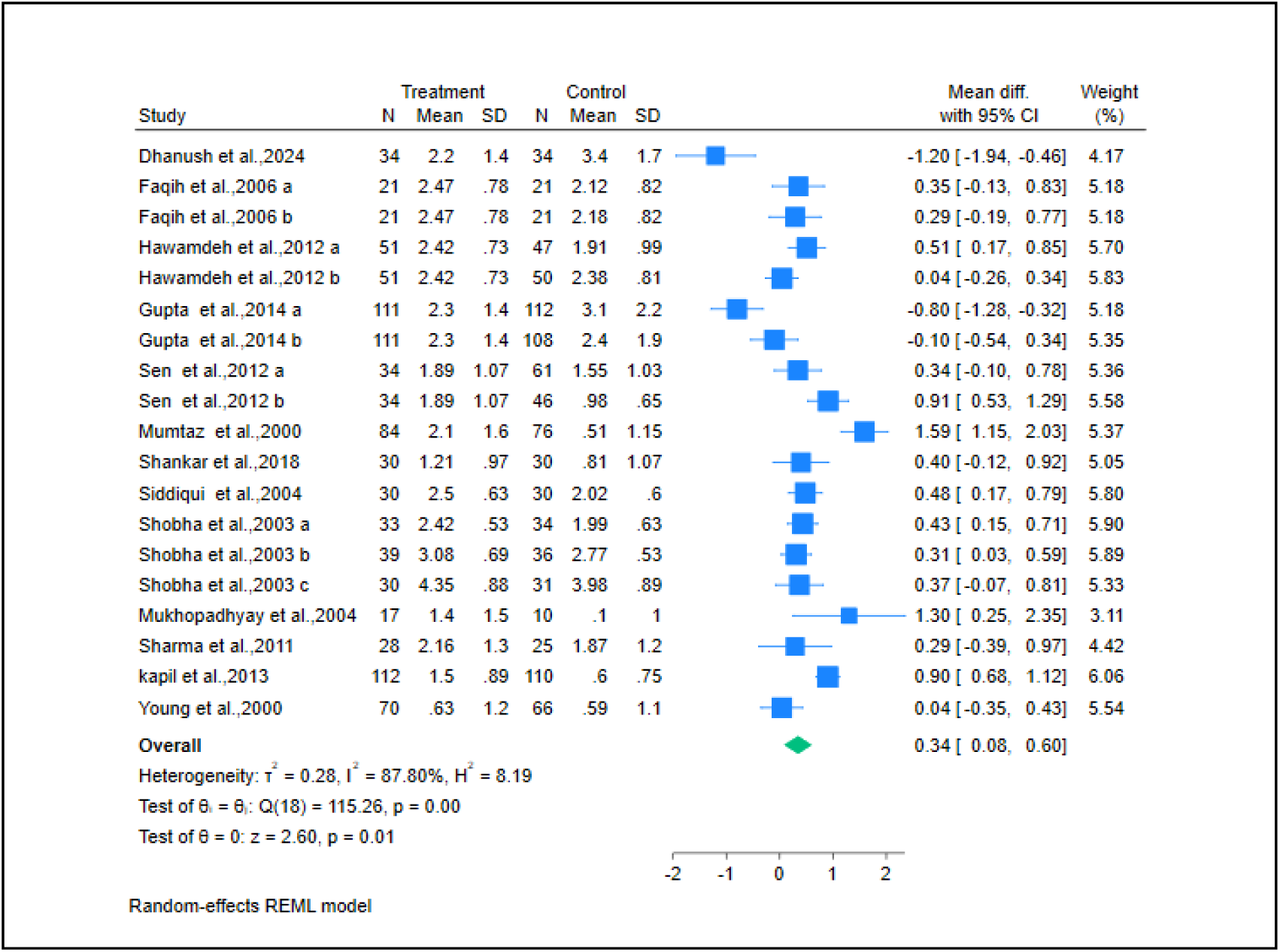
Forest plot showing the pooled mean difference in haemoglobin levels (g/dL) between daily and intermittent iron supplementation. **\*Treatment = Daily, *Control = Intermittent**

The mean (SD) end-of-intervention haemoglobin concentration across the included studies was 11.73 (0.64) g/dL in the daily supplementation group and 11.60 (0.87) g/dL in the intermittent supplementation group. The study by Kaundal et al. was excluded from the quantitative synthesis because haemoglobin outcomes were reported as medians without corresponding means and standard deviations. Nevertheless, its findings were consistent with the overall direction of the pooled estimate. Specifically, the study reported significantly greater median increases in haemoglobin in the daily supplementation group than in the alternate-day supplementation group at both 3 weeks (1.6 vs 1.1 g/dL; p = 0.02) and 6 weeks (2.9 vs 2.0 g/dL; p = 0.03).

### Subgroup analysis by population

Results showed that increase in haemoglobin levels was higher in daily supplementation than intermittent supplementation among children (MD: 0.44 g/dL, 95% CI: 0.19, 0.68; I² = 68.1%; n = 242 in the daily group (DG) vs n = 304 in the intermittent group (IG)). No notable difference in the pooled effect size was noted when a non-randomized study by Sharma et al., was excluded from the analysis (MD: 0.45 g/dL, 95% CI: 0.19, 0.71; I² = 72.8%). A similar direction of effect was observed among pregnant women (MD: 0.79 g/dL, 95% CI: 0.04, 1.55; I² = 87.8%; n = 201 DG vs n = 182 IG, respectively). In contrast, no statistically significant difference was observed among adolescent girls (MD: 0.22 g/dL, 95% CI: -0.16, 0.60; I² = 86.0%; n = 247 DG and n = 428 IG, respectively). Following exclusion of the non-randomized study by Sen et al., the effect estimate was attenuated and remained non-significant (MD: 0.07 g/dL, 95% CI: -0.37, 0.50; I² = 85.4%).

Among women of reproductive age, only one eligible study was available, which showed a higher increase in haemoglobin levels with intermittent supplementation than with daily supplementation (MD: -1.20 g/dL, 95% CI: -1.94, -0.46; n = 34 in each group). Because this subgroup included only one study, the finding should be interpreted with caution.

The test for subgroup differences was statistically significant (Q = 18.81; p < 0.001), indicating that the effect of daily versus intermittent supplementation differed across participant populations (Figure 3).

**Figure 3:**
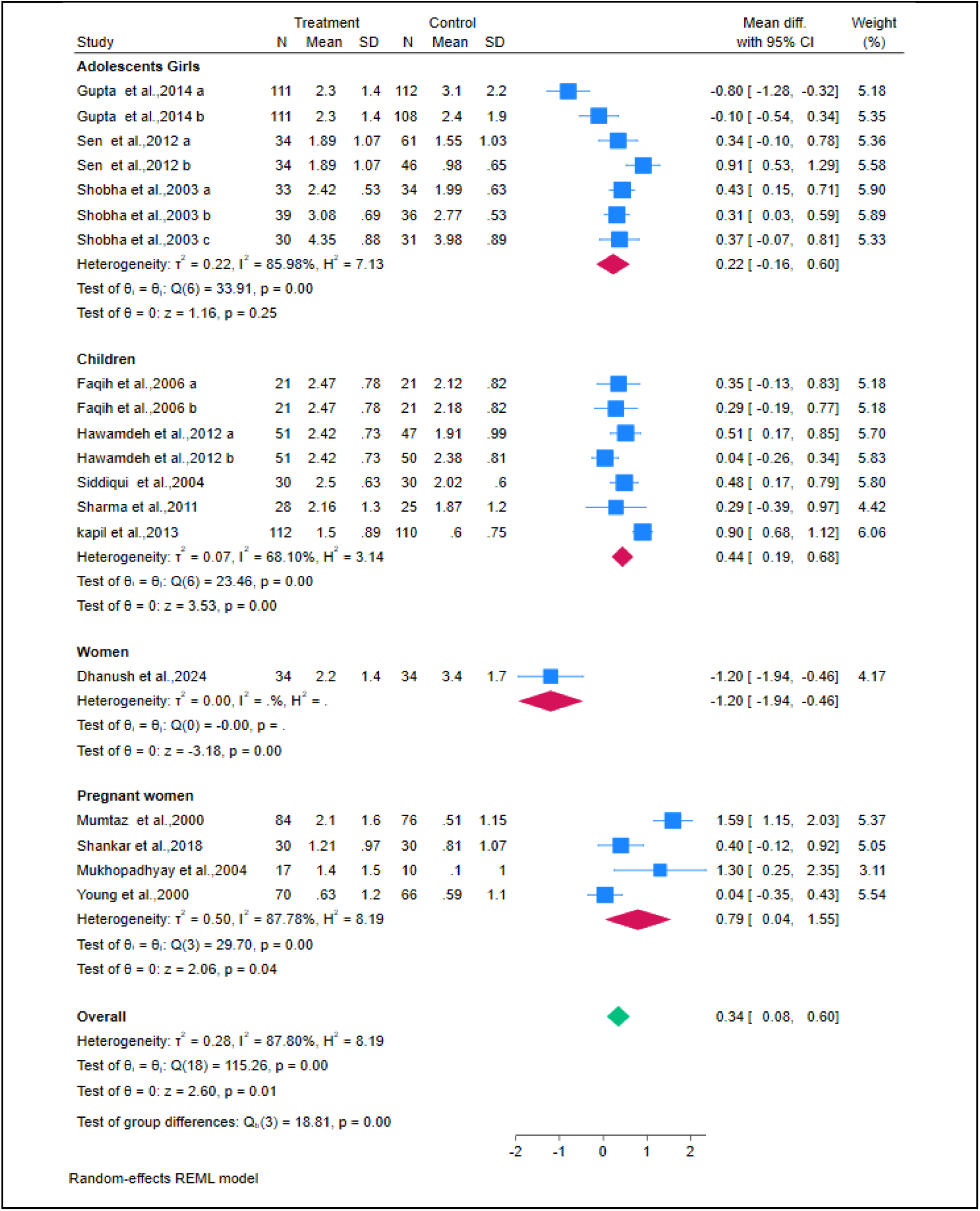
Forest plot of subgroup analyses according to population type. **\*Treatment = Daily, *Control = Intermittent**

### Serum ferritin

The meta-analysis of serum ferritin concentrations included five studies contributing seven comparisons, comprising 276 participants receiving daily supplementation and 398 receiving intermittent supplementation. The pooled mean difference (MD) was 4.48 µg/L (95% CI 0.37 to 8.59; I² = 52.4%), favouring daily supplementation. Daily iron supplementation was associated with a small but statistically significant increase in serum ferritin concentration compared with intermittent regimens (Z = 2.14; p = 0.03). Moderate between-study heterogeneity was observed (I² = 52.4%), indicating some variability in treatment effects across studies. Individual study findings were heterogeneous. Significant increases in serum ferritin were reported in two studies (Gupta et al. and Mumtaz et al.), whereas the remaining studies did not demonstrate statistically significant differences between daily and intermittent supplementation. Despite this variability, the pooled estimate consistently favoured daily supplementation (Figure 4).

**Figure 4:**
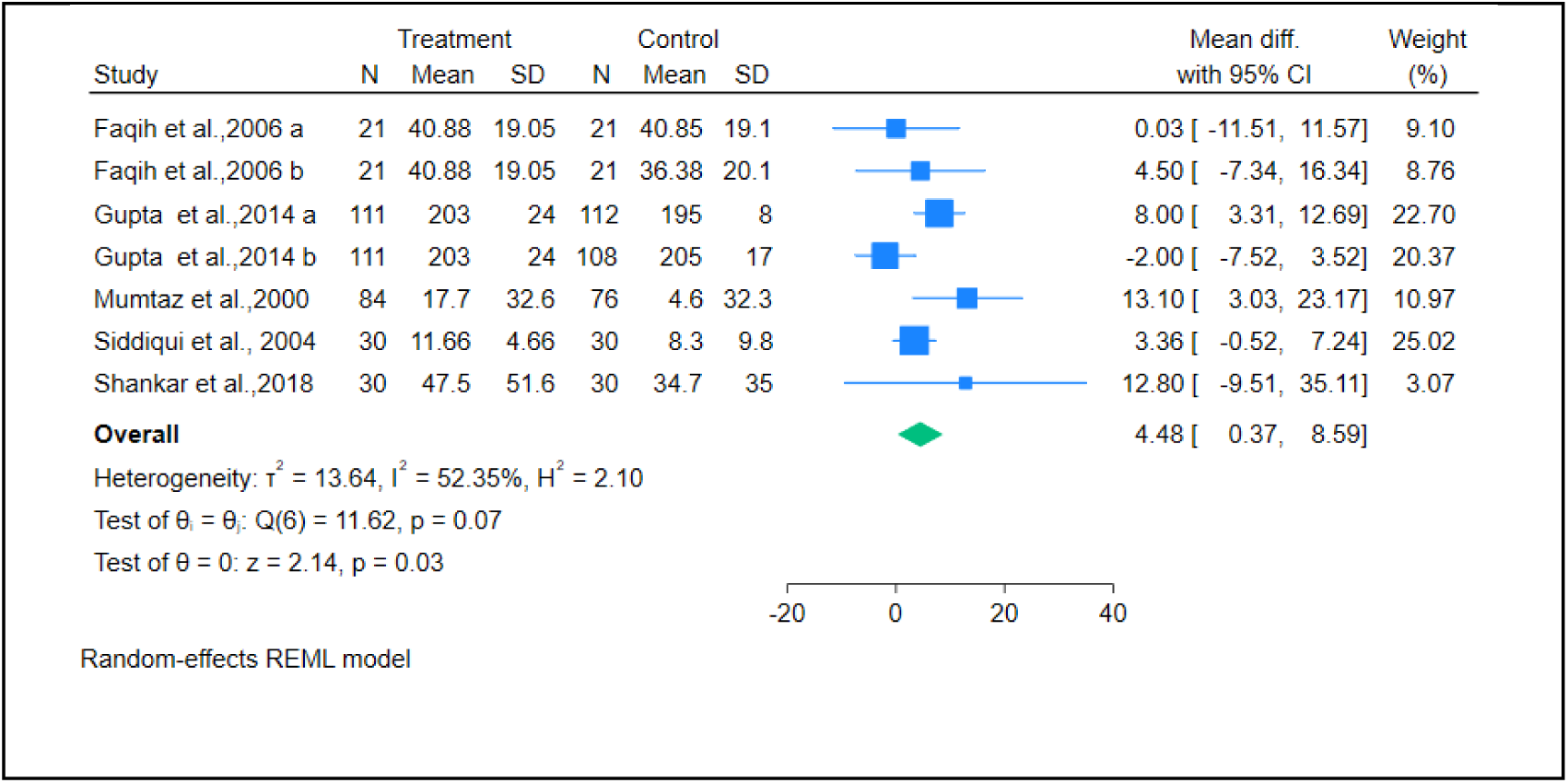
Forest plot pooled mean difference in serum ferritin levels (μg/L) between daily and intermittent iron supplementation. **\*Treatment = Daily, *Control = Intermittent**

### Secondary outcomes Recovery from anaemia

Six studies reported outcomes related to recovery from anaemia. Among young children, findings were inconsistent, with most studies reporting comparable recovery rates between daily and intermittent regimens. Sharma et al. observed similar reductions in anaemia prevalence with daily and weekly supplementation (35), while Hawamdeh et al. and Faqih et al. found no statistically significant differences in anaemia resolution between dosing schedules (25,27). In contrast, Kapil et al. reported higher recovery with daily supplementation than with weekly supplementation among preschool children (36). Among adolescent girls, both studies reported comparable recovery irrespective of dosing frequency. Shobha and Sharada et al. observed normalization of haemoglobin among all participants with mild or moderate anaemia in both treatment groups (32), while Sen and Kanani et al. reported similar haemoglobin improvements after one year of supplementation with daily and twice-weekly regimens (30).

### Maternal and fetal outcomes

Only two studies reported feto-maternal data. Mukhopadhyay and colleagues reported no significant differences in birth weight (daily: 2.9 ± 0.3 kg, weekly: 2.9 ± 0.4 kg), low birth weight incidence (daily: 10% vs weekly: 12.5%), or gestational age at delivery (daily: 38.6 ± 1.4, weekly: 38.3 ± 1.3 weeks) between groups. Shankar et al. reported that among anemic primigravida women (Hb 8-11 g/dL) receiving daily versus weekly iron folic acid tablets until six weeks postpartum, gestational length (daily: 275.1 ± 3.1 days; weekly: 274.5 ± 3.1 days), infant weight (daily: 2913.7 ± 62.6 g; weekly: 2864.7 ± 75.5 g), and placental weight (daily: 507.0 ± 19.3 g; weekly: 506.8 ± 24.5 g) were comparable between supplementation groups, with no significant differences observed (28,31).

### Adherence

Six studies assessed adherence or treatment compliance. Overall, intermittent supplementation was associated with better adherence than daily supplementation across most studies.

Among pregnant women, Mukhopadhyay et al. reported substantially higher adherence with weekly supplementation than with daily supplementation, with gastrointestinal adverse effects accounting for most treatment discontinuation in the daily group (28). Young et al. similarly observed higher adherence with weekly supplementation and fewer side effects (34).

Among adolescent girls, Gupta et al. and Shobha and Sharada et al. also reported better adherence with intermittent regimens, accompanied by substantially fewer adverse effects (26,32). In contrast, Dhanush et al. found similarly high adherence in both daily and alternate-day groups (24), while Kaundal et al. reported that no participants discontinued treatment because of adverse events (37).

### Adverse effects of iron supplementation

Five studies reported adverse events, which were predominantly gastrointestinal, including nausea, vomiting, epigastric discomfort, constipation, and diarrhoea. Across all studies, adverse events occurred more frequently with daily supplementation than with intermittent regimens.

Among non-pregnant women, Dhanush et al. and Kaundal et al. both reported significantly fewer gastrointestinal adverse effects with alternate-day supplementation than with daily dosing (24,37). Among pregnant women, Mukhopadhyay et al. observed a substantially lower frequency of gastrointestinal side effects with weekly supplementation than with daily supplementation (28). Among adolescent girls, Gupta et al. and Shobha and Sharada et al. consistently reported markedly lower rates of adverse effects among participants receiving intermittent supplementation (26,32). Overall, the evidence consistently demonstrated that intermittent iron supplementation was associated with a lower burden of gastrointestinal adverse effects than daily supplementation.

### Long-term outcomes

None of the included studies reported long-term clinical outcomes. Outcome assessment was limited to the end of supplementation or, in studies of pregnant women, up to six weeks postpartum, with no post-intervention follow-up.

### Risk of bias

The methodological quality of the included studies is summarised in Figures 5 and 6. Among randomised trials, most studies appeared to have a low risk of bias, particularly for outcome measurement and completeness of outcome data. Several studies raised some concerns related to the randomization process and lack of blinding, while Faqih et al. demonstrated a higher risk of bias in domains related to allocation and adherence. Kaundal et al., included only in the narrative synthesis, was assessed as having a low risk of bias across all RoB 2 domains. The two non-randomised studies, assessed using the ROBINS-I tool, seemed to have an overall moderate risk of bias, primarily due to confounding and selection of reported results, whereas risks related to intervention classification and outcome measurement were low. Overall, the methodological quality of the included evidence was considered acceptable for inclusion in the review and meta-analysis (Figures 5 and 6).

**Figure 5a and 5b:**
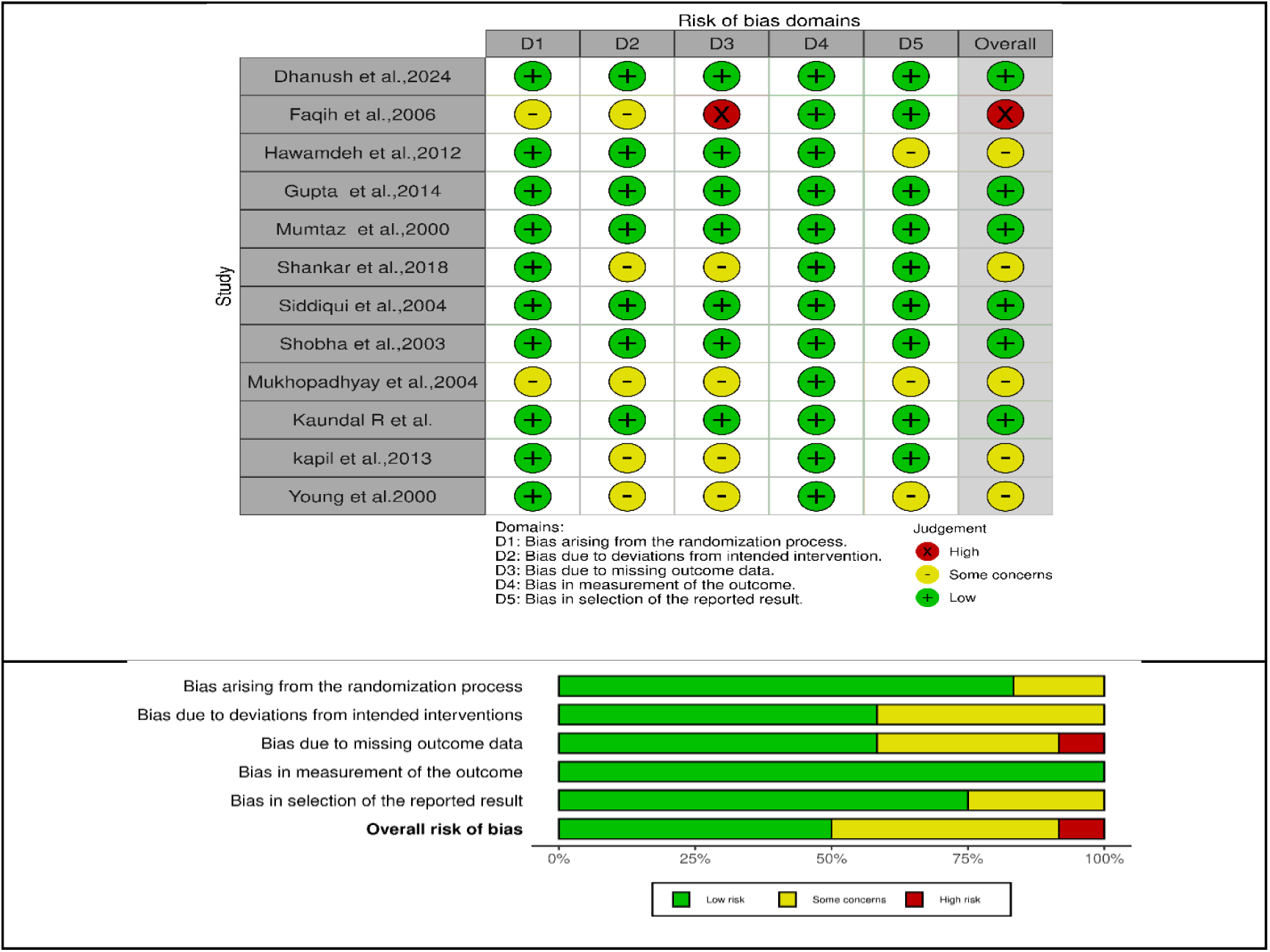
Risk of bias assessment for the included RCTs using the Cochrane Risk of Bias 2.0 (RoB 2) tool.

**Figure 6a and 6b:**
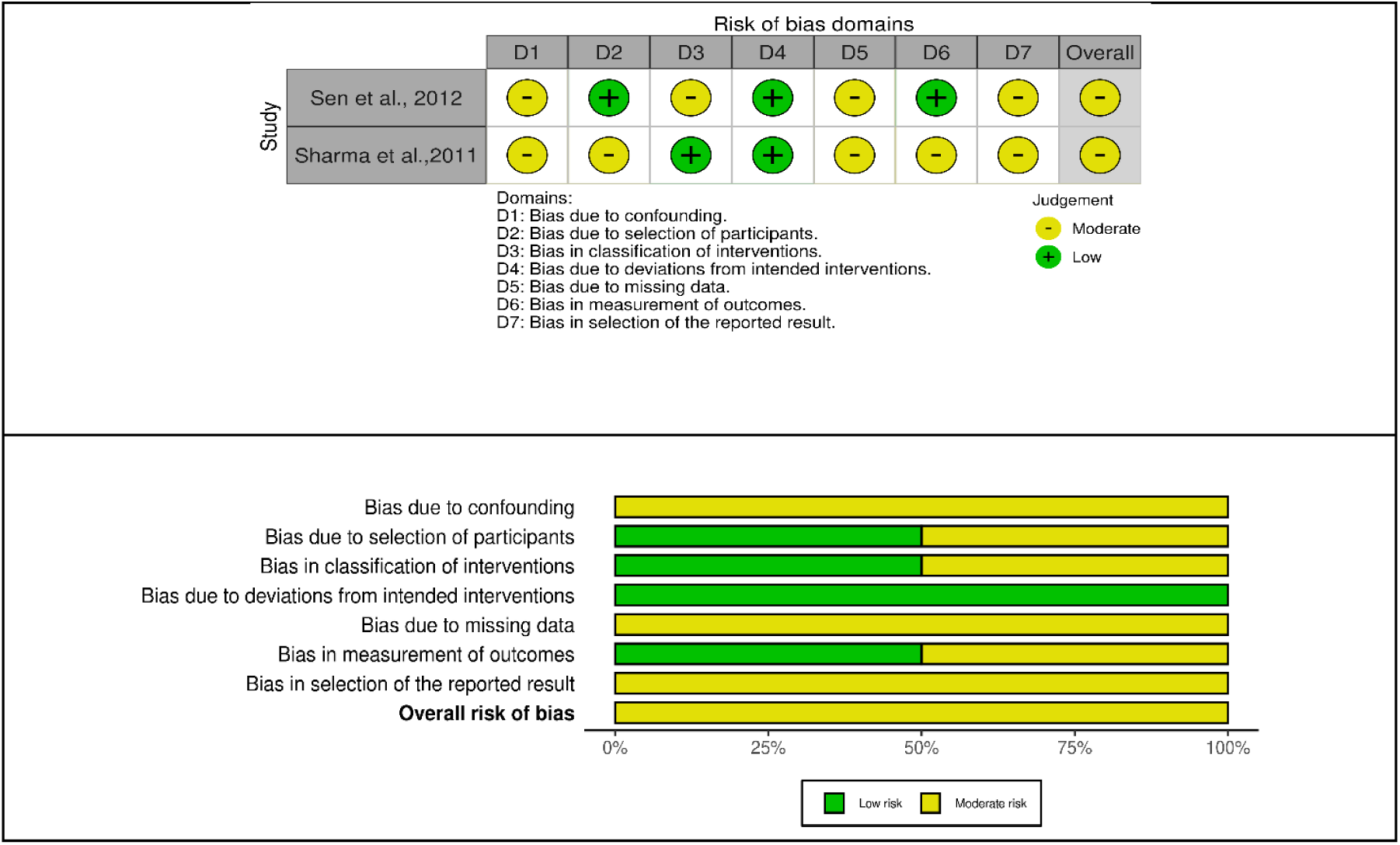
Risk of bias assessment for non-randomised studies using the ROBINS-I tool.

### Publication bias

Publication bias for the primary outcome of change in haemoglobin concentration was assessed using visual inspection of the funnel plot and Egger’s regression test for small-study effects. The funnel plot (Figure 7), including 13 studies (19 comparisons), appeared visually symmetrical. Consistent with the visual assessment, Egger’s regression test showed no evidence of small-study effects or publication bias (Z = - 0.60; p = 0.54).

**Figure 7:**
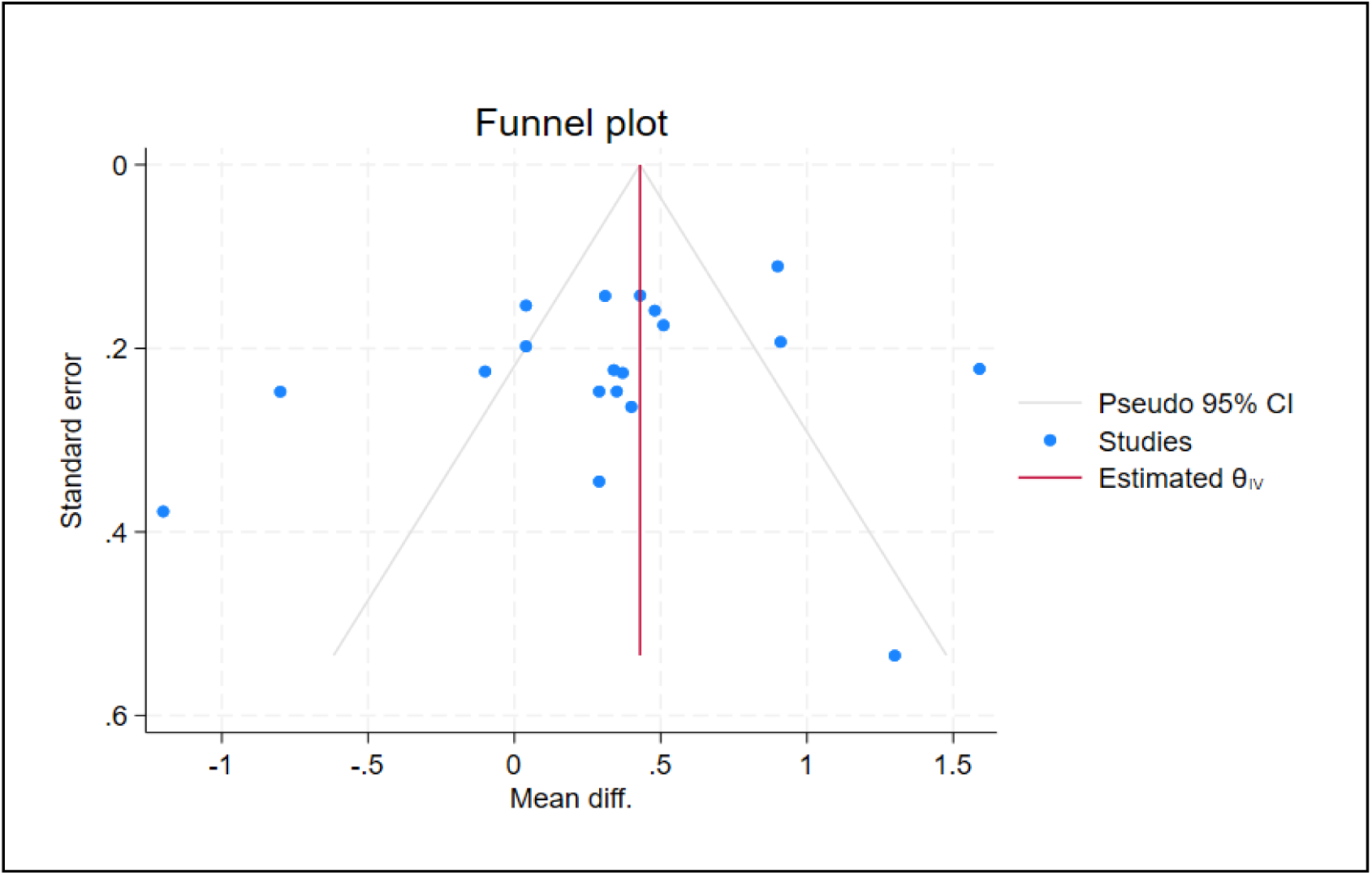
Funnel plot of included studies for haemoglobin change.

### Certainty of evidence

The certainty of evidence for the primary outcomes was assessed using the GRADE approach and is summarised in the Summary of Findings table (Supplementary Table 3).

The certainty of evidence for both haemoglobin change and serum ferritin concentration was rated as very low, primarily because of substantial statistical heterogeneity and methodological limitations, including unclear allocation concealment and lack of blinding in several included trials.

### Sensitivity analysis

Sensitivity analysis was conducted to assess the robustness of the pooled effect estimate for haemoglobin change. Across all iterations, the pooled mean difference remained stable, ranging from 0.28 to 0.41 g/dL. No individual study had a disproportionate influence on the overall pooled estimate, indicating that the primary findings were robust and were not driven by any single study or statistical outlier (Supplementary Figure 1).

### Meta-regression

Random-effects meta-regression was performed to explore potential sources of heterogeneity between the studies. Sample size (N) (β = -0.001, p = 0.50) and supplementation duration (β = - 0.02, p = 0.49) were not significantly associated with the intervention effect. Population category was also not a significant moderator overall, although the coefficient for non-pregnant women differed significantly from the reference category (β = -1.42, p = 0.02), however, it was represented by only one study. Supplementation regimen frequency was a significant moderator, with both twice-weekly (β = 1.52, p = 0.01) and weekly (β = 1.68, p = 0.006) regimens showing significantly different effect estimates compared with the daily regimen. These findings should be interpreted with caution because of the limited number of included studies (Supplementary Table 2).

## Discussion

In this systematic review and meta-analysis of 14 studies conducted in low- and middle-income countries (LMICs), we found that daily oral iron (or iron-folic acid) supplementation resulted in a statistically significant but modest improvement in haemoglobin concentration and serum ferritin compared with intermittent regimens among anaemic children, adolescents, women of reproductive age, and pregnant women. However, the absolute magnitude of benefit was small (0.34 g/dL for haemoglobin and 4.48 µg/L for serum ferritin), and the certainty of evidence was very low because of substantial heterogeneity and methodological limitations across the included studies. Importantly, intermittent supplementation consistently demonstrated better treatment adherence and fewer gastrointestinal adverse effects, while evidence for anaemia recovery and maternal or fetal outcomes was largely comparable between dosing schedules.

Our findings are broadly consistent with previous systematic reviews comparing different oral iron dosing schedules regarding treatment of anaemia in LMICs. Dhanvijay et al. reported a similarly modest advantage of daily over alternate-day supplementation for haemoglobin improvement (MD 0.28 g/dL; 95% CI −0.01 to 0.56), although heterogeneity remained substantial (17). Likewise, Kamath et al. concluded that alternate-day iron therapy achieved similar haematological responses while consistently reducing gastrointestinal adverse effects (18). However, both reviews largely excluded children, pregnant women, or weekly supplementation regimens, thereby limiting their applicability to LMIC populations with higher burden of anaemia. Our review addresses this important gap by synthesising evidence across multiple vulnerable populations and evaluating alternate-day, weekly and twice-weekly dosing schedules within a single analysis.

The modest superiority of daily supplementation is biologically plausible and supported by current understanding of iron metabolism. Oral iron administration induces a transient increase in circulating hepcidin concentrations, which suppresses intestinal iron absorption for approximately 24 hours (15). Consequently, alternate-day or intermittent dosing permits hepcidin concentrations to decline before subsequent doses, improving fractional iron absorption (16). Nevertheless, despite lower absorption efficiency per dose, daily supplementation provides a higher cumulative iron intake over time, which likely explains the modest but consistent improvements in haemoglobin and ferritin observed in this review.

The significant subgroup differences observed across populations suggest that physiological iron requirements influence treatment response. Children experienced the greatest benefit from daily supplementation, consistent with the Cochrane review by De-Regil et al., which concluded that daily supplementation was more effective than intermittent regimens for improving haemoglobin and reducing anaemia in children (19). Similarly, Andersen et al., in a meta-analysis of 129 randomised trials, demonstrated that more frequent supplementation schedules (three to seven doses per week) resulted in larger improvements in haemoglobin and serum ferritin than once- or twice-weekly regimens (38). In contrast, no significant benefit of daily supplementation was observed among adolescents, possibly owing to greater biological variability related to pubertal growth, menstrual blood loss, dietary habits, and differences in adherence across studies (39).

Our findings have important implications for anaemia control programmes in LMICs. WHO currently recommends daily oral iron supplementation for treatment of iron deficiency anaemia and for routine supplementation among high-risk groups (9,10). Nevertheless, programme implementation frequently encounters poor adherence due to gastrointestinal intolerance and prolonged treatment duration (11,12). Our findings support maintaining daily supplementation as the preferred first-line regimen, particularly in patients with moderate or severe anaemia requiring rapid correction. However, intermittent regimens may represent a pragmatic alternative for individuals who are unable to tolerate daily therapy or who repeatedly demonstrate poor adherence.

This review has several strengths. To our knowledge, it is one of the first systematic review to evaluate daily versus intermittent oral iron supplementation specifically for the treatment of anaemia across multiple vulnerable populations exclusively in LMICs. The review used validated risk-of-bias tools, assessed certainty of evidence using the GRADE framework, and demonstrated robust findings through sensitivity analyses with no evidence of publication bias. Restricting the review to LMICs also improves the applicability of the findings to settings where anaemia prevalence is highest and health-system constraints frequently influence treatment adherence.

Several limitations should also be acknowledged. First, substantial between-study heterogeneity persisted despite subgroup analyses, reflecting variation in participant characteristics, iron formulations, elemental iron doses, supplementation frequency and intervention duration. Second, the certainty of evidence for the primary outcomes was rated as very low because several studies had methodological limitations, including inadequate reporting of randomisation procedures, lack of blinding and relatively small sample sizes. Third, important clinical outcomes including anaemia recovery, functional status, cognitive performance, quality of life and long-term maternal and neonatal outcomes were infrequently reported, limiting assessment of whether modest improvements in haemoglobin translate into clinically meaningful benefits. Fourth, evidence among non-pregnant women of reproductive age was limited to a single study, preventing robust conclusions for this important population. Finally, the small number of studies evaluating alternate-day, weekly and twice-weekly regimens separately precluded regimen-specific meta-analyses that may have identified the optimal intermittent dosing strategy.

Future research should prioritise adequately powered randomised controlled trials comparing alternate-day, weekly and twice-weekly supplementation using standardised elemental iron doses and clinically relevant outcomes. Future studies should evaluate anaemia resolution, functional capacity, cognitive performance, maternal and neonatal outcomes, treatment adherence, quality of life and cost-effectiveness. In addition, mechanistic studies incorporating biomarkers of iron metabolism, including hepcidin, may help identify patient groups that derive the greatest benefit from different dosing schedules. Such evidence will facilitate development of more personalised iron supplementation strategies that optimise both efficacy and tolerability.

In conclusion, daily oral iron supplementation provides modest improvements in haemoglobin concentration and iron stores than intermittent regimens among anaemic populations in LMICs. However, these benefits are small, supported by very low-certainty evidence, and should be interpreted alongside the consistently superior adherence and lower gastrointestinal adverse effects associated with intermittent therapy. Daily supplementation should remain the preferred treatment for iron deficiency anaemia, particularly where rapid correction is clinically indicated, whereas intermittent regimens represent a reasonable alternative for individuals unable to tolerate or adhere to daily treatment.

## Data Availability

All data analysed in this study are derived from previously published studies and are included in this manuscript and its supplementary materials. The original studies are available through the cited references and the bibliographic databases from which they were retrieved. No new datasets were generated during this study.

## OSF REGISTRATION

https://doi.org/10.17605/OSF.IO/EFWTV

## Declarations

### Ethics approval and consent to participate

Not applicable. This review synthesised data from published and publicly available sources. No ethical approval was required as only aggregate data were used.

### Consent for publication

Not applicable.

### Availability of data and materials

All data supporting the findings of this review are included within the article and its supplementary files. Additional data are available from the corresponding author upon reasonable request.

### Authors’ contributions

Pratibha Dhiman drafted the manuscript. Krushna Chandra Sahoo and Subhanwita Manna developed search strategy. Pratibha Dhiman, Mrunali Zode, Ranadip Chowdhary, Pravin Kumar, Devoleena Chatterjee and Arshi Ahad contributed to study screening, data extraction, analysis, and manuscript preparation. Reema Mukherjee and Tanica Lyngdoh provided overall supervision, methodological guidance, and critically reviewed the manuscript. All authors read and approved the final manuscript.

