## supplementary material for "Daily Versus Intermittent Oral Iron Supplementation for the Treatment of Anaemia in Low- and Middle-Income Countries: A Systematic Review and Meta-analysis"

### PRISMA 2020 Checklist

| Section and Topic | Item # | Checklist item | Location where item is reported |
|---|---|---|---|
| **TITLE** | | | |
| Title | 1 | Identify the report as a systematic review. | **Page 1**, Title (Lines **1-2**) |
| **ABSTRACT** | | | |
| Abstract | 2 | See the PRISMA 2020 for Abstracts checklist. | **Pages 2-3** (Lines **29-66**) |
| **INTRODUCTION** | | | |
| Rationale | 3 | Describe the rationale for the review in the context of existing knowledge. | **Pages 3-4**, Lines 84-**119** |
| Objectives | 4 | Provide an explicit statement of the objective(s) or question(s) the review addresses. | **Pages 4-5**, Lines 120-**126** |
| **METHODS** | | | |
| Eligibility criteria | 5 | Specify the inclusion and exclusion criteria for the review and how studies were grouped for the syntheses. | **Page 5**, Lines **134-146** |
| Information sources | 6 | Specify all databases, registers, websites, organisations, reference lists and other sources searched or consulted to identify studies. Specify the date when each source was last searched or consulted. | **Page 5-6**, Lines **147-156** |
| Search strategy | 7 | Present the full search strategies for all databases, registers and websites, including any filters and limits used. | **Page 5-6**, **Supplementary Table 1** |
| Selection process | 8 | Specify the methods used to decide whether a study met the inclusion criteria of the review, including how many reviewers screened each record and each report retrieved, whether they worked independently, and if applicable, details of automation tools used in the process. | **Page 6**, Lines **157-162** |
| Data collection process | 9 | Specify the methods used to collect data from reports, including how many reviewers collected data from each report, whether they worked independently, any processes for obtaining or confirming data from study investigators, and if applicable, details of automation tools used in the process. | **Page 6**, Lines **176-186** |
| Data items | 10a | List and define all outcomes for which data were sought. Specify whether all results that were compatible with each outcome domain in each study were sought (e.g. for all measures, time points, analyses), and if not, the methods used to decide which results to collect. | **Page 6**, Lines **163-175** |
| | 10b | List and define all other variables for which data were sought (e.g. participant and intervention characteristics, funding sources). Describe any assumptions made about any missing or unclear information. | **Page 6**, Lines **163-175** |
| Study risk of bias assessment | 11 | Specify the methods used to assess risk of bias in the included studies, including details of the tool(s) used, how many reviewers assessed each study and whether they worked independently, and if applicable, details of automation tools used in the process. | **Page 7**, Lines **188-192** |
| Effect measures | 12 | Specify for each outcome the effect measure(s) (e.g. risk ratio, mean difference) used in the synthesis or presentation of results. | **Page 7** |
| Synthesis methods | 13a | Describe the processes used to decide which studies were eligible for each synthesis (e.g. tabulating the study intervention characteristics and comparing against the planned groups for each synthesis (item #5)). | **Page 5-7**, Lines **128-207** |
| | 13b | Describe any methods required to prepare the data for presentation or synthesis, such as handling of missing summary statistics, or data conversions. | **Page 6- 7**, Lines **176-207** |
| | 13c | Describe any methods used to tabulate or visually display results of individual studies and syntheses. | **Page 8-18**, Tables and Figures (Table 1; Figures 1-7) |

#### PRISMA 2020 Checklist

| Section and Topic | Item # | Checklist item | Location where item is reported |
|---|---|---|---|
| | 13d | Describe any methods used to synthesize results and provide a rationale for the choice(s). If meta-analysis was performed, describe the model(s), method(s) to identify the presence and extent of statistical heterogeneity, and software package(s) used. | **Page 7,** Lines **194-207** |
| | 13e | Describe any methods used to explore possible causes of heterogeneity among study results (e.g. subgroup analysis, meta-regression). | **Page 7,** Lines **193-207** |
| | 13f | Describe any sensitivity analyses conducted to assess robustness of the synthesized results. | **Page 7,** Lines 204-207 |
| Reporting bias assessment | 14 | Describe any methods used to assess risk of bias due to missing results in a synthesis (arising from reporting biases). | **Page 7,** Lines 187-192 |
| Certainty assessment | 15 | Describe any methods used to assess certainty (or confidence) in the body of evidence for an outcome. | **Page 7,** Lines 206-207 |
| **RESULTS** | | | |
| Study selection | 16a | Describe the results of the search and selection process, from the number of records identified in the search to the number of studies included in the review, ideally using a flow diagram. | **Page 8-9,** Lines 214-243 |
| | 16b | Cite studies that might appear to meet the inclusion criteria, but which were excluded, and explain why they were excluded. | **Page 8,** figure-1 |
| Study characteristics | 17 | Cite each included study and present its characteristics. | **Page 8-11, Table 1** |
| Risk of bias in studies | 18 | Present assessments of risk of bias for each included study. | **Page 15, figure 5&6** |
| Results of individual studies | 19 | For all outcomes, present, for each study: (a) summary statistics for each group (where appropriate) and (b) an effect estimates and its precision (e.g. confidence/credible interval), ideally using structured tables or plots. | **Page 8-15, Table 1, Figure 2-4** |
| Results of syntheses | 20a | For each synthesis, briefly summarise the characteristics and risk of bias among contributing studies. | Pages 9-18 |
| | 20b | Present results of all statistical syntheses conducted. If meta-analysis was done, present for each the summary estimate and its precision (e.g. confidence/credible interval) and measures of statistical heterogeneity. If comparing groups, describe the direction of the effect. | **Pages 9- 18**, Figures 2-4 |
| | 20c | Present results of all investigations of possible causes of heterogeneity among study results. | **Pages 12-13**, subgroup analysis |
| | 20d | Present results of all sensitivity analyses conducted to assess the robustness of the synthesized results. | **Pages 17, 30**, **Supplementary Figure 1** |
| Reporting biases | 21 | Present assessments of risk of bias due to missing results (arising from reporting biases) for each synthesis assessed. | **Pages 15-17** |
| Certainty of evidence | 22 | Present assessments of certainty (or confidence) in the body of evidence for each outcome assessed. | **Pages 17** |
| **DISCUSSION** | | | |
| Discussion | 23a | Provide a general interpretation of the results in the context of other evidence. | **Pages 18-20** |
| | 23b | Discuss any limitations of the evidence included in the review. | **Pages 18-20, Lines 470-482)** |
| | 23c | Discuss any limitations of the review processes used. | **Pages 19** |

### PRISMA 2020 Checklist

| Section and Topic | Item # | Checklist item | Location where item is reported |
|---|---|---|---|
| | 23d | Discuss implications of the results for practice, policy, and future research. | **Pages 19-20 Lines 492-499** |
| **OTHER INFORMATION** | | | |
| Registration and protocol | 24a | Provide registration information for the review, including register name and registration number, or state that the review was not registered. | **Page 5**, Lines 128-130, OSF Registration |
| | 24b | Indicate where the review protocol can be accessed, or state that a protocol was not prepared. | **Page 5** |
| | 24c | Describe and explain any amendments to information provided at registration or in the protocol. | NA |
| Support | 25 | Describe sources of financial or non-financial support for the review, and the role of the funders or sponsors in the review. | NA |
| Competing interests | 26 | Declare any competing interests of review authors. | **Page 20** |
| Availability of data, code and other materials | 27 | Report which of the following are publicly available and where they can be found: template data collection forms; data extracted from included studies; data used for all analyses; analytic code; any other materials used in the review. | **Page 20** |

*From:* Page MJ, McKenzie JE, Bossuyt PM, Boutron I, Hoffmann TC, Mulrow CD, et al. The PRISMA 2020 statement: an updated guideline for reporting systematic reviews. BMJ 2021;372:n71. doi: 10.1136/bmj.n71. This work is licensed under CC BY 4.0. To view a copy of this license, visit https://creativecommons.org/licenses/by/4.0/

**Supplementary Table 1: Search Strategy**

| | |
|---|---|
| **Pubmed (n= 1107)** | ("Iron"[MeSH Terms] OR "Folic Acid"[MeSH Terms] OR "iron-folic acid"[Title/Abstract] OR "iron-folic acid"[Title/Abstract] OR "IFA"[Title/Abstract] OR "iron supplementation"[Title/Abstract] OR "folic acid supplementation"[Title/Abstract] OR "iron and folic acid supplementation"[Title/Abstract] OR "iron-folate"[Title/Abstract] OR "iron-folate supplementation"[Title/Abstract] OR "micronutrient supplementation"[Title/Abstract] OR "iron tablet"[Title/Abstract] OR "folic acid tablet"[Title/Abstract]) AND ("Drug Administration Schedule"[MeSH Terms] OR "daily"[Title/Abstract] OR "every day"[Title/Abstract] OR "once daily"[Title/Abstract] OR "continuous"[Title/Abstract] OR "regular"[Title/Abstract] OR "intermittent"[Title/Abstract] OR "non-daily"[Title/Abstract] OR "alternate day"[Title/Abstract] OR "weekly"[Title/Abstract] OR "twice weekly"[Title/Abstract] OR "dosing schedule"[Title/Abstract] OR "frequency"[Title/Abstract]) AND ("Haemoglobins"[MeSH Terms] OR "Anaemia"[MeSH Terms] OR "haemoglobin"[Title/Abstract] OR "haemoglobin"[Title/Abstract] OR "haemoglobin concentration"[Title/Abstract] OR "Anaemia"[Title/Abstract] OR "anaemia"[Title/Abstract] OR "iron deficiency"[Title/Abstract] OR "serum ferritin"[Title/Abstract] OR "ferritin"[Title/Abstract] OR "hematologic"[Title/Abstract] OR "haematologic"[Title/Abstract] OR "blood cell count"[Title/Abstract] OR "RBC count"[Title/Abstract] OR "erythrocyte indices"[Title/Abstract] OR "hematological parameter*"[Title/Abstract] OR "haematological parameter*"[Title/Abstract] OR "Red Blood Cell Count"[Title/Abstract] OR "Hematologic Diseases"[Title/Abstract]) AND ("Vulnerable Populations"[MeSH Terms] OR "child"[Title/Abstract] OR "children"[Title/Abstract] OR "infant*"[Title/Abstract] OR "baby"[Title/Abstract] OR "babies"[Title/Abstract] OR "adolescent*"[Title/Abstract] OR "teenager*"[Title/Abstract] OR "youth"[Title/Abstract] OR "young people"[Title/Abstract] OR "women of reproductive age"[Title/Abstract] OR "Reproductive age women"[Title/Abstract] OR "WRA"[Title/Abstract] OR "preconception women"[Title/Abstract] OR "pregnant"[Title/Abstract] OR "pregnancy"[Title/Abstract] OR "pregnant women"[Title/Abstract] OR "expecting mothers"[Title/Abstract] OR "mother*"[Title/Abstract] OR "female*"[Title/Abstract] OR "girls"[Title/Abstract] OR "vulnerable population*"[Title/Abstract] OR "high risk group*"[Title/Abstract] OR "at risk population*"[Title/Abstract])AND ("Developing Countries"[MeSH Terms] OR "Africa"[MeSH Terms] OR "Asia"[MeSH Terms] OR "South America"[MeSH Terms] OR "Latin America"[MeSH Terms] OR "Africa South of the Sahara"[MeSH Terms] OR "asia, southeastern"[MeSH Terms] OR "asia, western"[MeSH Terms] OR "Caribbean Region"[MeSH Terms] OR "Central America"[MeSH Terms] OR "Pacific Islands"[MeSH Terms] OR "LMIC"[Title/Abstract] OR "LMICs"[Title/Abstract] OR "low and middle income countr*"[Title/Abstract] OR "developing countr*"[Title/Abstract] OR "resource-limited setting"[Title/Abstract] OR "resource-poor setting"[Title/Abstract] OR "resource-poor settings"[Title/Abstract] OR "low income countr*"[Title/Abstract] OR "middle income countr*"[Title/Abstract] OR "underdeveloped countr*"[Title/Abstract] OR "less developed countr*"[Title/Abstract] OR "least developed countr*"[Title/Abstract] OR "low resource setting"[Title/Abstract] OR "low resource settings"[Title/Abstract] OR "transitional countr*"[Title/Abstract] OR "economically developing countr*"[Title/Abstract] OR ("Afghanistan"[All Fields] OR "Albania"[All Fields] OR "Algeria"[All Fields] OR "American Samoa"[All Fields] OR "Angola"[All Fields] OR "Argentina"[All Fields] OR "Armenia"[All Fields] OR "Azerbaijan"[All Fields] OR "Bangladesh"[All Fields] OR "Belarus"[All Fields] OR "Belize"[All Fields] OR "Benin"[All Fields] OR "Bhutan"[All Fields] OR "Bolivia"[All Fields] OR "Bosnia and Herzegovina"[All Fields] OR "Botswana"[All Fields] OR "Brazil"[All Fields] OR "Bulgaria"[All Fields] OR "Burkina Faso"[All Fields] OR "Burundi"[All Fields] OR "Cabo Verde"[All Fields] OR "Cambodia"[All Fields] OR "Cameroon"[All Fields] OR "Central African Republic"[All Fields] OR "Chad"[All Fields] OR "China"[All Fields] OR "Colombia"[All Fields] OR "Comoros"[All Fields] OR "Congo"[All Fields] OR (("Congo"[MeSH Terms] OR "Congo"[All Fields]) |

| | |
|---|---|
| | AND "dem"[All Fields] AND "rep"[All Fields]) OR (("Congo"[MeSH Terms] OR "Congo"[All Fields]) AND "rep"[All Fields]) OR "Costa Rica"[All Fields] OR "Cote d'Ivoire"[All Fields] OR "Croatia"[All Fields] OR "Cuba"[All Fields] OR "Dominica"[All Fields] OR "Dominican Republic"[All Fields] OR "Djibouti"[All Fields] OR "Ecuador"[All Fields] OR "Egypt"[All Fields] OR (("Egypt"[MeSH Terms] OR "Egypt"[All Fields] OR "egypt s"[All Fields]) AND ("arabs"[MeSH Terms] OR "arabs"[All Fields] OR "arab"[All Fields]) AND "rep"[All Fields]) OR "El Salvador"[All Fields] OR "Equatorial Guinea"[All Fields] OR "Eritrea"[All Fields] OR "Eswatini"[All Fields] OR "Ethiopia"[All Fields] OR "Fiji"[All Fields] OR "Gabon"[All Fields] OR "Gambia"[All Fields] OR "Georgia"[All Fields] OR "Ghana"[All Fields] OR "Grenada"[All Fields] OR "Guatemala"[All Fields] OR "Guinea"[All Fields] OR "Guinea-Bissau"[All Fields] OR "Guyana"[All Fields] OR "Honduras"[All Fields] OR "Hungary"[All Fields] OR "India"[All Fields] OR "Indonesia"[All Fields] OR "Iran"[All Fields] OR "Iraq"[All Fields] OR "Jamaica"[All Fields] OR "Jordan"[All Fields] OR "Kazakhstan"[All Fields] OR "Kenya"[All Fields] OR "Kiribati"[All Fields] OR (("korea"[MeSH Terms] OR "korea"[All Fields] OR "korea s"[All Fields] OR "koreas"[All Fields]) AND "dem"[All Fields] AND ("people s"[All Fields] OR "peopled"[All Fields] OR "peopling"[All Fields] OR "persons"[MeSH Terms] OR "persons"[All Fields] OR "people"[All Fields] OR "peoples"[All Fields]) AND "rep"[All Fields]) OR "Kosovo"[All Fields] OR "Kyrgyz Republic"[All Fields] OR "Lao PDR"[All Fields] OR "Lebanon"[All Fields] OR "Lesotho"[All Fields] OR "Liberia"[All Fields] OR "Libya"[All Fields] OR "Madagascar"[All Fields] OR "Malawi"[All Fields] OR "Malaysia"[All Fields] OR "Maldives"[All Fields] OR "Mali"[All Fields] OR "Marshall Islands"[All Fields] OR "Mauritania"[All Fields] OR "Mauritius"[All Fields] OR "Mexico"[All Fields] OR (("micronesia"[MeSH Terms] OR "micronesia"[All Fields]) AND "fed"[All Fields] AND "sts"[All Fields]) OR "Moldova"[All Fields] OR "Mongolia"[All Fields] OR "Montenegro"[All Fields] OR "Morocco"[All Fields] OR "Mozambique"[All Fields] OR "Myanmar"[All Fields] OR "Namibia"[All Fields] OR "Nepal"[All Fields] OR "Nicaragua"[All Fields] OR "Niger"[All Fields] OR "Nigeria"[All Fields] OR "North Macedonia"[All Fields] OR "Pakistan"[All Fields] OR "Palau"[All Fields] OR "Panama"[All Fields] OR "Papua New Guinea"[All Fields] OR "Paraguay"[All Fields] OR "Peru"[All Fields] OR "Philippines"[All Fields] OR "Russian Federation"[All Fields] OR "Rwanda"[All Fields] OR "Samoa"[All Fields] OR "Sao Tome and Principe"[All Fields] OR "Senegal"[All Fields] OR "Serbia"[All Fields] OR "Seychelles"[All Fields] OR "Sierra Leone"[All Fields] OR "Solomon Islands"[All Fields] OR "Somalia"[All Fields] OR "South Africa"[All Fields] OR "South Sudan"[All Fields] OR "Sri Lanka"[All Fields] OR "st lucia"[All Fields] OR "st vincent and the grenadines"[All Fields] OR "Sudan"[All Fields] OR "Suriname"[All Fields] OR "Syrian Arab Republic"[All Fields] OR "Tajikistan"[All Fields] OR "Tanzania"[All Fields] OR "Thailand"[All Fields] OR "Timor-Leste"[All Fields] OR "Togo"[All Fields] OR "Tonga"[All Fields] OR "Trinidad and Tobago"[All Fields] OR "Tunisia"[All Fields] OR "Turkey"[All Fields] OR "Turkmenistan"[All Fields] OR "Tuvalu"[All Fields] OR "Uganda"[All Fields] OR "Ukraine"[All Fields] OR "Uruguay"[All Fields] OR "Uzbekistan"[All Fields] OR "Vanuatu"[All Fields] OR "Venezuela"[All Fields] OR (("Venezuela"[MeSH Terms] OR "Venezuela"[All Fields] OR "venezuela s"[All Fields]) AND "rb"[All Fields]) OR "Vietnam"[All Fields] OR "Zambia"[All Fields] OR "Zimbabwe"[All Fields])) |
| **Web of science (n= 1065)** | (Iron OR "Folic Acid" OR "iron-folic acid" OR "iron-folic acid" OR IFA OR "iron supplementation" OR "folic acid supplementation" OR "iron and folic acid supplementation" OR iron-folate OR "iron-folate supplementation" OR "micronutrient supplementation" OR "iron tablet" OR "folic acid tablet") AND ("Drug Administration Schedule" OR daily OR "every day" OR "once daily" OR continuous OR regular OR intermittent OR non-daily OR "alternate day" OR weekly OR "twice weekly" OR "dosing schedule" OR frequency) AND (Haemoglobins OR Anaemia OR haemoglobin OR haemoglobin OR "haemoglobin concentration" OR Anaemia OR anaemia OR "iron deficiency" OR "serum ferritin" OR ferritin OR hematologic OR haematologic OR "blood cell count" OR "RBC count" OR "erythrocyte indices" OR "hematological parameter*" OR "haematological parameter*" OR "Red Blood Cell Count" OR "Hematologic |

| | |
|---|---|
| | Diseases") AND ("Vulnerable Populations" OR child OR children OR infant* OR baby OR babies OR adolescent* OR teenager* OR youth OR "young people" OR "women of reproductive age" OR "Reproductive age women" OR WRA OR "preconception women" OR pregnant OR pregnancy OR "pregnant women" OR "expecting mothers" OR mother* OR female* OR girls OR "vulnerable population*" OR "high risk group*" OR "at risk population*") AND("Developing Countries" OR Africa OR Asia OR "South America" OR "Latin America" OR "Africa South of the Sahara" OR "asia, southeastern" OR "asia, western" OR "Caribbean Region" OR "Central America" OR "Pacific Islands" OR LMIC OR LMICs OR "low and middle income countr*" OR "developing countr*" OR "resource-limited setting" OR "resource-poor setting" OR "resource-poor settings" OR "low income countr*" OR "middle income countr*" OR "underdeveloped countr*" OR "less developed countr*" OR "least developed countr*" OR "low resource setting" OR "low resource settings" OR "transitional countr*" OR "economically developing countr*" OR (Afghanistan OR Albania OR Algeria OR "American Samoa" OR Angola OR Argentina OR Armenia OR Azerbaijan OR Bangladesh OR Belarus OR Belize OR Benin OR Bhutan OR Bolivia OR "Bosnia and Herzegovina" OR Botswana OR Brazil OR Bulgaria OR "Burkina Faso" OR Burundi OR "Cabo Verde" OR Cambodia OR Cameroon OR "Central African Republic" OR Chad OR China OR Colombia OR Comoros OR Congo OR ((Congo OR Congo) AND dem AND rep) OR ((Congo OR Congo) AND rep) OR "Costa Rica" OR "Cote d'Ivoire" OR Croatia OR Cuba OR Dominica OR "Dominican Republic" OR Djibouti OR Ecuador OR Egypt OR ((Egypt OR Egypt OR "egypt s") AND (arabs OR arabs OR arab) AND rep) OR "El Salvador" OR "Equatorial Guinea" OR Eritrea OR Eswatini OR Ethiopia OR Fiji OR Gabon OR Gambia OR Georgia OR Ghana OR Grenada OR Guatemala OR Guinea OR Guinea-Bissau OR Guyana OR Honduras OR Hungary OR India OR Indonesia OR Iran OR Iraq OR Jamaica OR Jordan OR Kazakhstan OR Kenya OR Kiribati OR ((korea OR korea OR "korea s" OR koreas) AND dem AND ("people s" OR peopled OR peopling OR persons OR persons OR people OR peoples) AND rep) OR Kosovo OR "Kyrgyz Republic" OR "Lao PDR" OR Lebanon OR Lesotho OR Liberia OR Libya OR Madagascar OR Malawi OR Malaysia OR Maldives OR Mali OR "Marshall Islands" OR Mauritania OR Mauritius OR Mexico OR ((micronesia OR micronesia) AND fed AND sts) OR Moldova OR Mongolia OR Montenegro OR Morocco OR Mozambique OR Myanmar OR Namibia OR Nepal OR Nicaragua OR Niger OR Nigeria OR "North Macedonia" OR Pakistan OR Palau OR Panama OR "Papua New Guinea" OR Paraguay OR Peru OR Philippines OR "Russian Federation" OR Rwanda OR Samoa OR "Sao Tome and Principe" OR Senegal OR Serbia OR Seychelles OR "Sierra Leone" OR "Solomon Islands" OR Somalia OR "South Africa" OR "South Sudan" OR "Sri Lanka" OR "st lucia" OR "st vincent and the grenadines" OR Sudan OR Suriname OR "Syrian Arab Republic" OR Tajikistan OR Tanzania OR Thailand OR Timor-Leste OR Togo OR Tonga OR "Trinidad and Tobago" OR Tunisia OR Turkey OR Turkmenistan OR Tuvalu OR Uganda OR Ukraine OR Uruguay OR Uzbekistan OR Vanuatu OR Venezuela OR ((Venezuela OR Venezuela OR "venezuela s") AND rb) OR Vietnam OR Zambia OR Zimbabwe)) |
| **Scopus (n=2624)** | (Iron OR "Folic Acid" OR "iron-folic acid" OR "iron-folic acid" OR IFA OR "iron supplementation" OR "folic acid supplementation" OR "iron and folic acid supplementation" OR iron-folate OR "iron-folate supplementation" OR "micronutrient supplementation" OR "iron tablet" OR "folic acid tablet") AND ("Drug Administration Schedule" OR daily OR "every day" OR "once daily" OR continuous OR regular OR intermittent OR non-daily OR "alternate day" OR weekly OR "twice weekly" OR "dosing schedule" OR frequency) AND (Haemoglobins OR Anaemia OR haemoglobin OR haemoglobin OR "haemoglobin concentration" OR Anaemia OR anaemia OR "iron deficiency" OR "serum ferritin" OR ferritin OR hematologic OR haematologic OR "blood cell count" OR "RBC count" OR "erythrocyte indices" OR "hematological parameter*" OR "haematological parameter*" OR "Red Blood Cell Count" OR "Hematologic Diseases") AND ("Vulnerable Populations" OR child OR children OR infant* OR baby OR babies OR adolescent* OR teenager* OR youth OR "young people" OR "women of reproductive age" OR "Reproductive age women" OR WRA OR "preconception women" OR pregnant OR pregnancy OR |

| | |
|---|---|
| | "pregnant women" OR "expecting mothers" OR mother* OR female* OR girls OR "vulnerable population*" OR "high risk group*" OR "at risk population*")<br>AND("Developing Countries" OR Africa OR Asia OR "South America" OR "Latin America" OR "Africa South of the Sahara" OR "asia, southeastern" OR "asia, western" OR "Caribbean Region" OR "Central America" OR "Pacific Islands" OR LMIC OR LMICs OR "low and middle income countr*" OR "developing countr*" OR "resource-limited setting" OR "resource-poor setting" OR "resource-poor settings" OR "low income countr*" OR "middle income countr*" OR "underdeveloped countr*" OR "less developed countr*" OR "least developed countr*" OR "low resource setting" OR "low resource settings" OR "transitional countr*" OR "economically developing countr*" OR (Afghanistan OR Albania OR Algeria OR "American Samoa" OR Angola OR Argentina OR Armenia OR Azerbaijan OR Bangladesh OR Belarus OR Belize OR Benin OR Bhutan OR Bolivia OR "Bosnia and Herzegovina" OR Botswana OR Brazil OR Bulgaria OR "Burkina Faso" OR Burundi OR "Cabo Verde" OR Cambodia OR Cameroon OR "Central African Republic" OR Chad OR China OR Colombia OR Comoros OR Congo OR ((Congo OR Congo) AND dem AND rep) OR ((Congo OR Congo) AND rep) OR "Costa Rica" OR "Cote d'Ivoire" OR Croatia OR Cuba OR Dominica OR "Dominican Republic" OR Djibouti OR Ecuador OR Egypt OR ((Egypt OR Egypt OR "egypt s") AND (arabs OR arabs OR arab) AND rep) OR "El Salvador" OR "Equatorial Guinea" OR Eritrea OR Eswatini OR Ethiopia OR Fiji OR Gabon OR Gambia OR Georgia OR Ghana OR Grenada OR Guatemala OR Guinea OR Guinea-Bissau OR Guyana OR Honduras OR Hungary OR India OR Indonesia OR Iran OR Iraq OR Jamaica OR Jordan OR Kazakhstan OR Kenya OR Kiribati OR ((korea OR korea OR "korea s" OR koreas) AND dem AND ("people s" OR peopled OR peopling OR persons OR persons OR people OR peoples) AND rep) OR Kosovo OR "Kyrgyz Republic" OR "Lao PDR" OR Lebanon OR Lesotho OR Liberia OR Libya OR Madagascar OR Malawi OR Malaysia OR Maldives OR Mali OR "Marshall Islands" OR Mauritania OR Mauritius OR Mexico OR ((micronesia OR micronesia) AND fed AND sts) OR Moldova OR Mongolia OR Montenegro OR Morocco OR Mozambique OR Myanmar OR Namibia OR Nepal OR Nicaragua OR Niger OR Nigeria OR "North Macedonia" OR Pakistan OR Palau OR Panama OR "Papua New Guinea" OR Paraguay OR Peru OR Philippines OR "Russian Federation" OR Rwanda OR Samoa OR "Sao Tome and Principe" OR Senegal OR Serbia OR Seychelles OR "Sierra Leone" OR "Solomon Islands" OR Somalia OR "South Africa" OR "South Sudan" OR "Sri Lanka" OR "st lucia" OR "st vincent and the grenadines" OR Sudan OR Suriname OR "Syrian Arab Republic" OR Tajikistan OR Tanzania OR Thailand OR Timor-Leste OR Togo OR Tonga OR "Trinidad and Tobago" OR Tunisia OR Turkey OR Turkmenistan OR Tuvalu OR Uganda OR Ukraine OR Uruguay OR Uzbekistan OR Vanuatu OR Venezuela OR ((Venezuela OR Venezuela OR "venezuela s") AND rb) OR Vietnam OR Zambia OR Zimbabwe)) |
| **Embase (n=1648)** | (Iron/exp OR 'Folic Acid'/exp OR 'iron-folic acid':ti,ab OR 'iron-folic acid':ti,ab OR IFA:ti,ab OR 'iron supplementation':ti,ab OR 'folic acid supplementation':ti,ab OR 'iron and folic acid supplementation':ti,ab OR iron-folate:ti,ab OR 'iron-folate supplementation':ti,ab OR 'micronutrient supplementation':ti,ab OR 'iron tablet':ti,ab OR 'folic acid tablet':ti,ab) AND ('Drug Administration Schedule'/exp OR daily:ti,ab OR 'every day':ti,ab OR 'once daily':ti,ab OR continuous:ti,ab OR regular:ti,ab OR intermittent:ti,ab OR non-daily:ti,ab OR 'alternate day':ti,ab OR weekly:ti,ab OR 'twice weekly':ti,ab OR 'dosing schedule':ti,ab OR frequency:ti,ab) AND (Haemoglobins/exp OR Anaemia/exp OR haemoglobin:ti,ab OR haemoglobin:ti,ab OR 'haemoglobin concentration':ti,ab OR Anaemia:ti,ab OR anaemia:ti,ab OR 'iron deficiency':ti,ab OR 'serum ferritin':ti,ab OR ferritin:ti,ab OR hematologic:ti,ab OR haematologic:ti,ab OR 'blood cell count':ti,ab OR 'RBC count':ti,ab OR 'erythrocyte indices':ti,ab OR 'hematological parameter*':ti,ab OR 'haematological parameter*':ti,ab OR 'Red Blood Cell Count':ti,ab OR 'Hematologic Diseases':ti,ab) AND ('Vulnerable Populations'/exp OR child:ti,ab OR children:ti,ab OR infant*:ti,ab OR baby:ti,ab OR babies:ti,ab OR adolescent*:ti,ab OR teenager*:ti,ab OR youth:ti,ab OR 'young people':ti,ab OR 'women of reproductive age':ti,ab OR 'Reproductive age women':ti,ab OR WRA:ti,ab OR 'preconception women':ti,ab OR pregnant:ti,ab OR pregnancy:ti,ab OR 'pregnant women':ti,ab OR 'expecting mothers':ti,ab OR mother*:ti,ab |

| | |
|---|---|
| | OR female*:ti,ab OR girls:ti,ab OR 'vulnerable population*':ti,ab OR 'high risk group*':ti,ab OR 'at risk population*':ti,ab)<br>AND<br><br>('Developing Countries'/exp OR Africa/exp OR Asia/exp OR 'South America'/exp OR 'Latin America'/exp OR 'Africa South of the Sahara'/exp OR 'asia, southeastern'/exp OR 'asia, western'/exp OR 'Caribbean Region'/exp OR 'Central America'/exp OR 'Pacific Islands'/exp OR LMIC:ti,ab OR LMICs:ti,ab OR 'low and middle income countr*':ti,ab OR 'developing countr*':ti,ab OR 'resource-limited setting':ti,ab OR 'resource-poor setting':ti,ab OR 'resource-poor settings':ti,ab OR 'low income countr*':ti,ab OR 'middle income countr*':ti,ab OR 'underdeveloped countr*':ti,ab OR 'less developed countr*':ti,ab OR 'least developed countr*':ti,ab OR 'low resource setting':ti,ab OR 'low resource settings':ti,ab OR 'transitional countr*':ti,ab OR 'economically developing countr*':ti,ab |

**Supplementary Table 2: Meta-regression analysis of sources of heterogeneity in the effect of iron supplementation**

| Moderator | | | | | z | p value | $\tau^2$ (residual) | $I^2$ % (residual) | $R^2$ % |
|---|---|---|---|---|---|---|---|---|---|
| | Coefficient | *SE* | 95% CI lower | 95% CI upper | *z* | *p* | $\tau^2$ | $I^2$ (%) | $R^2$ (%) |
| **Total Sample Size** | | | | | | | | | |
| N | -0.001 | 0.001 | -0.004 | 0.002 | 0.67 | 0.50 | 0.28 | 88.2 | 0.00 |
| *_con* | *0.50* | *0.28* | *- 0.043* | *1.05* | *1.81* | *0.07* | - | - | - |
| **Duration of supplementation** | | | | | | | | | |
| Duration (Month) | -0·02 | 0·03 | -0·09 | 0·04 | - 0·69 | 0·49 | 0·28 | 88.2% | 0·00% |
| *_cons* | 0·46 | 0·21 | -0·03 | 0·88 | 2·12 | 0·03 | - | - | - |
| **Population category** | | | | | | | | | |
| Children | 0·19 | 0·26 | -0·31 | 0·70 | 0·75 | 0·45 | - | - | - |
| Women (non-pregnant) | -1·42 | 0·61 | -2·62 | - 0·22 | - 2·34 | 0·02 | | - | - |
| Pregnant women | 0·54 | 0·32 | -0·09 | 1.17 | 1·67 | 0·09 | - | - | - |
| _cons (Adolescent) | 0·22 | 0·18 | -0·13 | 0·58 | 1·23 | 0·22 | 0·19 | 83·89% | 28.78% |
| **Control category (Intermittent)** | | | | | | | | | |
| Twice weekly | 1·52 | 0·61 | 0·31 | 2·74 | 2·73 | 0·01 | - | - | |
| Weekly | 1·68 | 0·61 | 0·47 | 2·88 | 2·74 | 0·006 | - | - | - |
| _cons (Daily) | -1·20 | 0·59 | -2·36 | - 0·38 | - 2·03 | 0·04 | 0·20 | 84.64% | 24.37% |

| Moderator | | | | | z | p value | $\tau^2$ (residual) | $I^2$ % (residual) | $R^2$ % |
|---|---|---|---|---|---|---|---|---|---|
| | Coefficient | *SE* | 95% CI lower | 95% CI upper | *z* | *p* | $\tau^2$ | $I^2$ (%) | $R^2$ (%) |

#### Supplementary Table 3: Certainty of Evidence Assessment (GRADE) for Haemoglobin and Ferritin Outcomes

**Summary of findings:**

Daily oral iron or iron-folic acid (IFA) supplementation compared to Intermittent oral iron or IFA supplementation for Anaemic children, adolescents, women of reproductive age, and pregnant women in LMICs

Patient or population: Anaemic children, adolescents, women of reproductive age, and pregnant women in LMICs

Setting: low- and middle-income countries (LMICs)

Intervention: Daily oral iron or iron-folic acid (IFA) supplementation

Comparison: Intermittent oral iron or IFA supplementation

| Outcomes | Anticipated absolute effects* (95% CI) | | Relative effect (95% CI) | № of participants (studies) | Certainty of the evidence (GRADE) | Comments |
|---|---|---|---|---|---|---|
| | Risk with Intermittent oral iron or IFA supplementation | Risk with Daily oral iron or iron-folic acid (IFA) supplementation | | | | |
| Haemoglobin change (Haemoglobin change) assessed with: g/dL | The mean haemoglobin change was 0 g/dL | MD 0.34g/dL higher (0.08 higher to 0.60higher) | - | 1672 (11 RCTs, two non-randomised studies) | ⊕◯◯◯ Very low[a,b,c] | Daily iron or iron-folic acid (IFA) supplementation may slightly increase haemoglobin levels compared to intermittent supplementation, but the effect is small and the certainty of evidence is very low. |
| Serum ferritin assessed with: µg/L | The mean new outcome was 0 µg/L | MD 4.48 µg/L higher (0.37 higher to 8.59 higher) | - | 674 (5 RCTs) | ⊕◯◯◯ Very low[ade] | Daily iron or iron-folic acid (IFA) supplementation may slightly increase serum ferritin levels compared to intermittent supplementation, but the confidence interval is wide and the certainty of evidence is very low |

**Summary of findings:**

Daily oral iron or iron-folic acid (IFA) supplementation compared to Intermittent oral iron or IFA supplementation for Anaemic children, adolescents, women of reproductive age, and pregnant women in LMICs

Patient or population: Anaemic children, adolescents, women of reproductive age, and pregnant women in LMICs

Setting: low- and middle-income countries (LMICs)

Intervention: Daily oral iron or iron-folic acid (IFA) supplementation

Comparison: Intermittent oral iron or IFA supplementation

| Outcomes | Anticipated absolute effects*(95% CI) | | Relative effect (95% CI) | № of participants (studies) | Certainty of the evidence (GRADE) | Comments |
|---|---|---|---|---|---|---|
| | Risk with Intermittent oral iron or IFA supplementation | Risk with Daily oral iron or iron-folic acid (IFA) supplementation | | | | |

*The risk in the intervention group (and its 95% confidence interval) is based on the assumed risk in the comparison group and the relative effect of the intervention (and its 95% CI).

CI: confidence interval; MD: mean difference

a. Downgraded one level for serious risk of bias due to concerns regarding randomization, allocation concealment, blinding, and incomplete outcome data in several included studies.

b. Downgraded one level for serious inconsistency because substantial statistical heterogeneity was observed for the haemoglobin outcome ($I^2$ = 87.8%).

c. Downgraded one level for serious indirectness because the pooled haemoglobin estimates combined different population groups (children, adolescents, women of reproductive age, and pregnant women).

d. Downgraded one level for serious inconsistency because moderate statistical heterogeneity was observed for the serum ferritin outcome ($I^2$ = 52.4%), with variability in effect estimates across the included studies.

e. Downgraded one level for serious imprecision because of the limited number of studies and wide confidence intervals.

GRADE Working Group grades of evidence
High certainty: we are very confident that the true effect lies close to that of the estimate of the effect.
Moderate certainty: we are moderately confident in the effect estimate: the true effect is likely to be close to the estimate of the effect, but there is a possibility that it is substantially different.
Low certainty: our confidence in the effect estimate is limited: the true effect may be substantially different from the estimate of the effect.
Very low certainty: we have very little confidence in the effect estimate: the true effect is likely to be substantially different from the estimate of effect.

**Supplementary Figure 1: Sensitivity analysis evaluating the robustness of pooled effect sizes for haemoglobin and serum ferritin.**

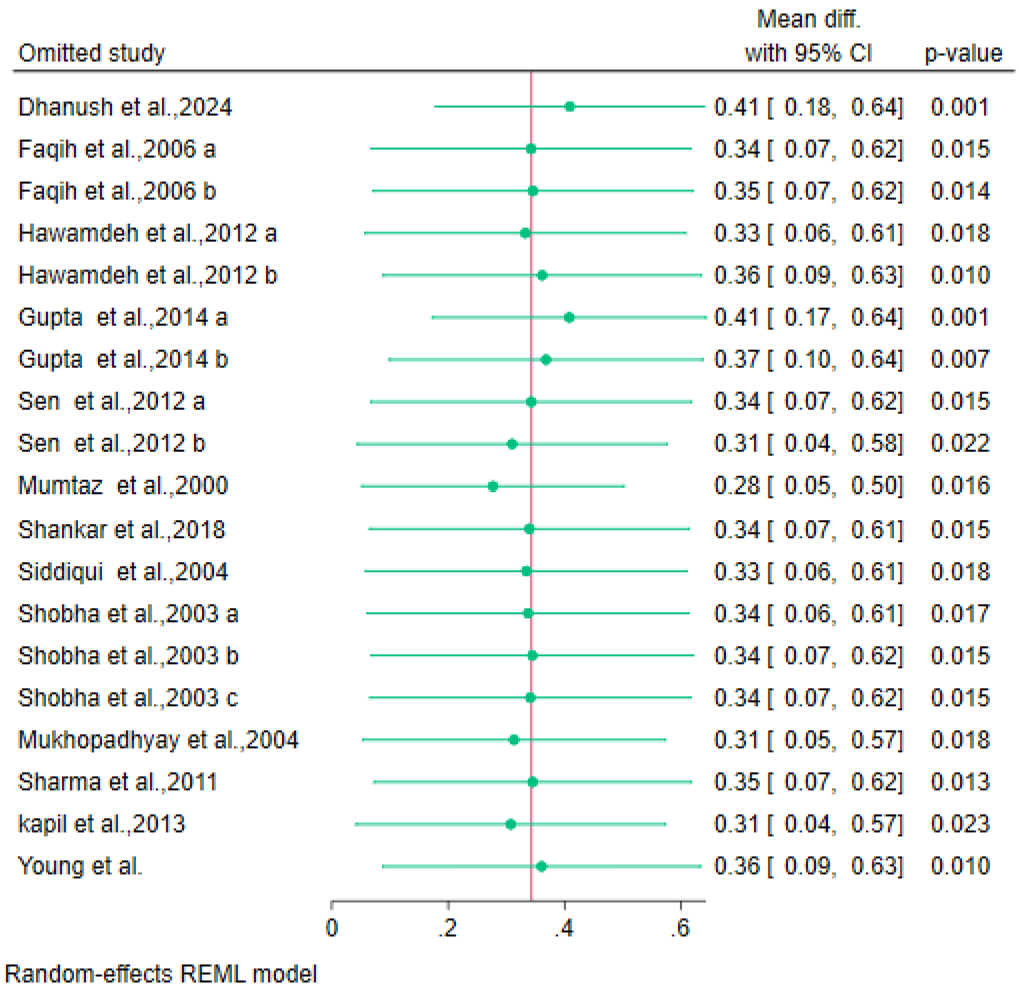
